# Admixture mapping identifies complex trait associations with local ancestry in the *All of Us Research Program*

**DOI:** 10.64898/2025.12.29.25343152

**Authors:** Wilfredo G Gonzalez Rivera, Youwen Liu, Tara Mirmira, Nichole Ma, Jihoon Kim, Matteo D’ Antonio, Umber Dube, Kelly A Frazer, Melissa Gymrek

**Affiliations:** Bioinformatics and Systems Biology Graduate Program, University of California, San Diego, La Jolla, CA, USA; Department of Medicine, Division of Biomedical Informatics, University of California, San Diego, La Jolla, CA, USA; Department of Biomedical Informatics & Data Science, Yale University, New Heaven, CT, USA; Department of Computer Science and Engineering, University of California San Diego, La Jolla, CA, USA; Department of Medicine, University of California San Diego, La Jolla, CA, USA; Medicine Diabetes Institute, University of Washington, Seattle, WA, USA; Department of Dermatology, University of California San Diego, La Jolla, CA, USA; Department of Pediatrics, University of California San Diego, La Jolla, California, USA; Institute of Genomic Medicine, University of California San Diego, La Jolla, California, USA

## Abstract

Genetic studies have largely focused on homogeneous populations, limiting our understanding of the genetic architecture of complex traits in admixed individuals. The advent of diverse biobanks like the *All of Us* Research Program (AoU) and computationally efficient local ancestry inference (LAI) methods now enable admixture mapping (ADM) at biobank scale. Here, we used two orthogonal LAI methods (GNOMIX and FLARE) to characterize local ancestry in the entire AoU v7.1 cohort (n=230,019). We then used GNOMIX labels to identify associations between African (AFR) and Native American (NAT) local ancestry with 29 quantitative traits. We first analyzed *All of Us* v7.1 data across African (*n*=49,797) and Admixed American (*n*=40,327) cohorts, which identified 97 significant local ancestry associations (65 AFR, 32 NAT). These include strong known signals, such as an association between AFR ancestry at the DARC locus and white blood cell traits and between NAT ancestry at the *BUD13/APOE5/ZPR1* locus and triglycerides, as well as additional signals not previously associated with local ancestry. We observed that trait associations with AFR local ancestry are largely consistent across the African and Admixed American cohorts, but that several AFR signals reach genome-wide significance exclusively in Admixed Americans. Grouping associations by trait category revealed distinct ancestral patterns: all endocrine, renal, and 75.0% of liver signals were driven by associations with NAT ancestry, whereas white blood cell (90.9%), red blood cell (65.1%), and lipid (66.7%) signals were largely associated with AFR ancestry, possibly reflecting different population-driven environmental exposures throughout history. Finally, we performed a second round of analysis, comprising the largest ADM study to date, on the entire AoU v7.1 cohort in which we pooled individuals from all ancestries. Despite evidence of confounding due to population structure, summary statistics for pooled results showed strong correlation (r>0.98) with those from single ancestry analysis and detected 2.7x fold more signals, most of which passed at least nominal significance and all of which showed consistent effect directions in the single ancestry cohorts. Overall, these results demonstrate the power of using large, admixed cohorts to gain new insights into the relationship between local ancestry and the genetic architecture of medically relevant complex traits.

## Introduction

A substantial fraction of the United States (U.S.) population is genetically admixed, meaning their genomes consist of segments from two or more ancestries. For example, 13.7% of US individuals self-identify as African American (with substantial African and European ancestry) and 19.5% self-identify as Hispanic/Latino (with substantial Native American, European, and African ancestry)^1^. These populations face a disproportionately high burden of certain complex diseases, many of which are strongly influenced by genetic factors. For example, liver disease ranks as one of the leading causes of death among Hispanic/Latino individuals, who have the highest prevalence of Metabolic Dysfunction-Associated Steatotic Liver Disease (MASLD)^2^ in the U.S. Similarly, cardiovascular disease disproportionately affects African American individuals^3^. However, these populations have been largely excluded from genetic studies, including genome-wide association studies (GWAS), which have mostly focused on homogeneous populations, particularly of European ancestry^4^. Addressing this inequity is essential for ensuring broad applicability of genomic studies across populations.

Admixture mapping (ADM) provides a powerful framework to study genetic contributions to disease in admixed individuals by testing for associations between local ancestry—defined as the ancestral origin of individual genomic segments—and complex traits^5,6^. ADM is particularly effective for identifying ancestry-specific variants that may be missed by standard GWAS methods and for identifying the ancestral background on which risk alleles are most common. Further, it may capture effects of other genetic variants that are missed by conventional GWAS, including structural variants, repeats, or haplotypes (combinations of variants in *cis*) that are tagged by local ancestry^7^. A classic example of a locus identified by ADM is the Duffy antigen receptor (*ACKR1*) locus, at which African local ancestry is strongly associated with lowered neutrophil count^8,9^. Many additional examples have been identified^10–13^. Despite its potential, ADM has been limited by small sample sizes of cohorts with admixed individuals, focus predominantly on two-way admixture (e.g. African vs. European ancestry in African Americans), and a lack of computational tools scalable to large datasets.

Recent methodological developments have substantially improved the scalability of local ancestry inference (LAI). RFMix^14^, which uses a random forest approach, is widely used but is computationally intensive, making it challenging to apply at biobank scale. More recent tools, such as FLARE^15^—a hidden Markov model-based method—and GNOMIX^16^—a supervised machine learning framework—offer substantial improvements in computational performance while maintaining high accuracy. At the same time, the availability of large-scale multi-ancestry cohorts such as the *All of Us Research Program*^17^ (AoU), which now includes nearly 500,000 genetically diverse participants from the U.S., offers a uniquely representative dataset to study the genetic architecture of complex traits in admixed populations. A recent study by Mandla et al.^18^ leveraged AoU to perform ADM study, but was limited to two-way admixture (African-European) and used only a subset of the AoU cohort (n=48,921).

In this study, we conducted the largest admixture mapping (ADM) analysis to date, leveraging data from the *All of Us* Research Program (v7.1) to identify associations between African (AFR) and Native American (NAT) ancestry and 29 quantitative traits. We first analyzed single-ancestry cohorts of African (*n*=49,797, considering AFR local ancestry) and Admixed American (*n*=40,327, considering NAT and AFR local ancestry) individuals. Results from these single-ancestry analyses were used to compare AFR-associated signals across cohorts and to identify traits with associations predominantly driven by either NAT or AFR local ancestry. Finally, we performed ADM analysis for NAT and AFR local ancestry across the entire AoU biobank and compared the results to those obtained based on single-ancestry analysis. Overall, our findings provide important insights into the genetic architecture of medically relevant traits—particularly within Hispanic/Latino and African American communities, who collectively comprise approximately one-third of the U.S. population and demonstrate the power of large multi-ancestry cohorts to identify ancestry-specific signals genetic associations.

## Results

### Local ancestry inference in the All of Us Research Program

We performed local ancestry inference (LAI) in the AoU v7.1 cohort using two complementary pipelines (**Fig. 1a**). The first, GNOMIX^16^, is a supervised machine learning method which assigns ancestry labels to a predefined set of 17,716 bins across the genome. The second, FLARE^15^, uses a hidden Markov model to infer the labels and boundaries of local ancestry segments. Both methods rely on a labeled set of reference haplotypes. For GNOMIX, we used their published model containing a total of 8 reference populations constructed primarily from the 1000Genomes^19^ and HGDP^20^ datasets. For FLARE, we constructed a custom reference haplotype panel containing a similar set of samples using 1000Genomes (European, East Asian, South Asian, and non-admixed African samples) and HGDP (Native Americans and Western Asians) individuals (total n=2,265). The full list of reference populations included in each label are provided in **Methods**. For comparison, we additionally performed global ancestry inference using two methods: ADMIXTURE^21^ and Rye^22^, in both cases using the custom reference panel constructed above.

**Figure 1:**
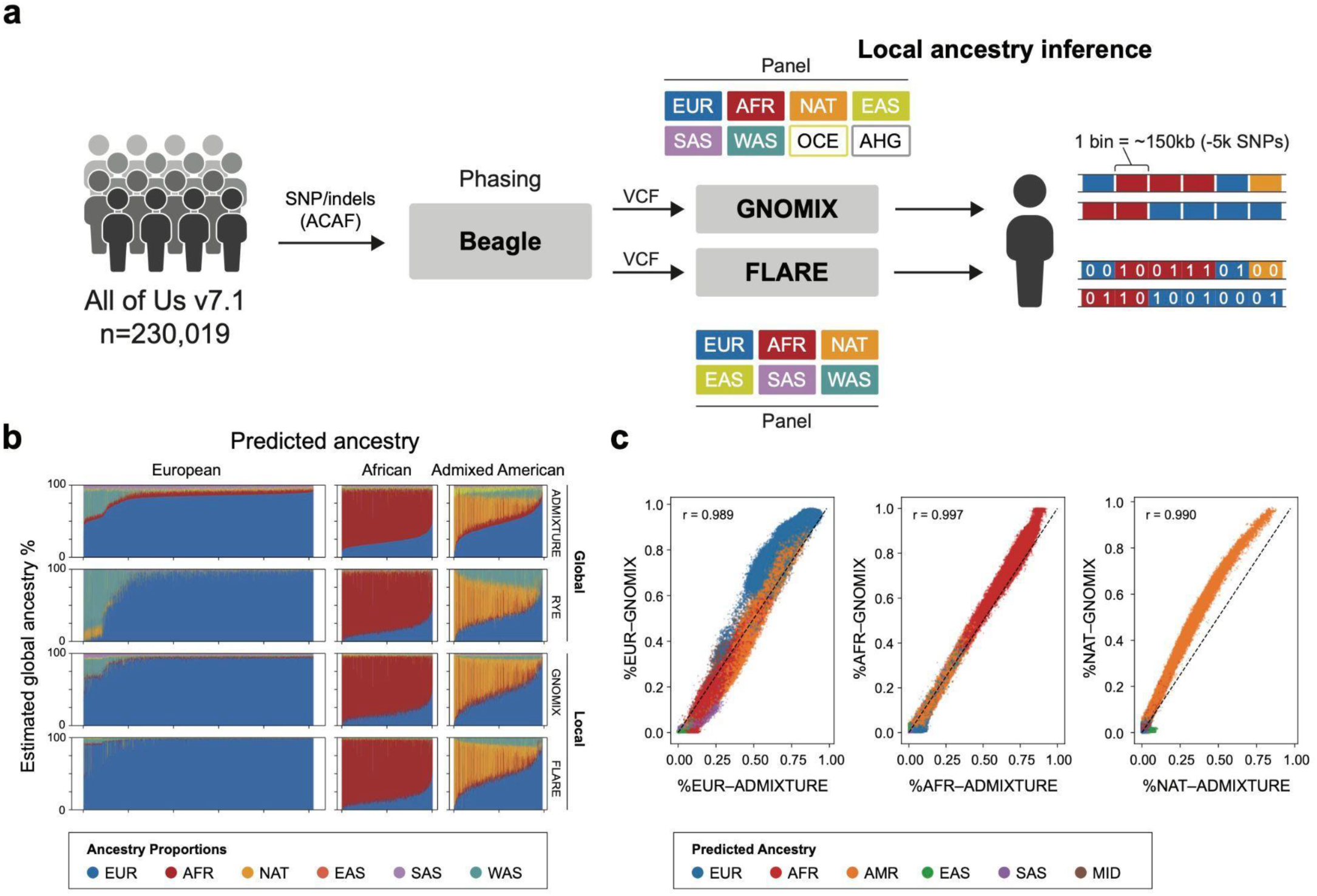
Local ancestry inference in the *All of Us Research Program*. **a. Schematic overview of local ancestry inference pipeline.** We applied two orthogonal pipelines for local ancestry inference (LAI). **Top:** LAI was performed using GNOMIX. GNOMIX outputs inferred labels for a predefined set of genomic windows. **Bottom:** LAI was performed using FLARE, which infers the local ancestry label for each input variant. Colored boxes denote the reference populations used by each tool (EUR=European; AFR=African; NAT=Native American; EAS=East Asian; SAS=South Asian; WAS=Western Asian; OCE=Oceanian; AHG=ancestral hunter gatherers). **b. Comparison of estimated global ancestry percentages stratified by predicted ancestry.** Plots show the percentages estimated using global ancestry inference (ADMIXTURE and Rye) and LAI (GNOMIX and FLARE). For GNOMIX and FLARE, estimates are based on the percent of the genome assigned to each label (**Methods**). 50,000 randomly chosen samples are shown. **c. Comparison of estimated global ancestry percentages for each predicted ancestry.** Plots show inferred global ancestry percentages based on ADMIXTURE (*x*-axis) and GNOMIX (*y*-axis) for each individual, colored by predicted ancestry labels provided by *All of Us*. Black lines give the x=y diagonal. Note, ADMIXTURE underestimates NAT compared to all other tools tested. A full pairwise comparison of all ancestry labels and the four tools is shown in **Fig. S1**.

To evaluate LAI results, we used several strategies. First, we reasoned that the percentage of the genome assigned to each local ancestry label based should be consistent with the global genetically predicted ancestry of each individual. Overall, we found results from all four tools recapitulated expected trends (**Fig. 1b; Table S1**). For example, self-reported White individuals whose genetically predicted ancestry is mostly European (EUR) showed predominantly European global ancestry (average estimates ranging 82.2%–91.3% across tools). For the Black and African Americans individuals, whose genetically predicted ancestry is mostly African (AFR) showed a mixture of African global ancestry (72.9%–81.0%) and European global ancestry (15.9%–21.7%). Lastly, for Hispanic and Latinos, whose genetically predicted ancestry is mostly Admixed Americans (AMR) demonstrated substantial admixture, with estimated European global ancestry (35.2% to 46.5%), Native American global ancestry (23.6% to 36.0%), African global ancestry (11.0% to 15.3%), and West Asian global ancestry (4.0% to 17.1%). We observed that Rye estimated a notably higher mean West Asian global ancestry in Admixed American individuals (17.1%) compared to other tools (range 4.0-8.0%), and likely reflects the consistent overestimation of West Asian ancestry observed across all groups using that method (see below).

Next, we compared the inferred percentages of comparable ancestry labels across all four methods. Percentages were highly correlated across methods (Pearson r > 0.94**, Fig. S1**) for all pairs of tools across five of the six mutual labels but showed some consistent biases across tools (**Fig. 1c; Fig. S1**). Most notably, estimates of West Asian and European ancestry in self-reported Middle Eastern or North African samples showed weaker correlation across methods, with FLARE estimates showing lowest overall concordance due to overestimating European ancestry and underestimating West Asian ancestry compared to the other tools. Further, compared to other methods, Rye consistently estimated higher percentages of West Asian ancestry in individuals from European predicted ancestry (**Fig. 1b; Fig. S1**). Our results are consistent with previous analyses by the AoU team, in which Rye estimated a substantial portion of West Asian ancestry in self-reported White individuals^17^. Finally, ADMIXTURE consistently underestimated NAT ancestry compared to all other tools (**Fig. 1c; Fig. S1**). Overall, our results suggest all four tools provide similar global ancestry estimates but demonstrate challenges in distinguishing between West Asian vs. European global ancestry.

### Admixture mapping in the Admixed American and African cohorts

We used inferred local ancestry labels to perform admixture mapping (ADM) for 29 quantitative traits based on commonly measured blood and serum biomarkers (**Fig. S2, Table S2**). We focused on GNOMIX labels for association testing (median window size 149kb; n=17,716 total windows), based on its high overall concordance with global ancestry estimates, high concordance with FLARE for AFR and NAT labels, and the lower computational burden compared to FLARE to test genomic windows rather than individual variants. Similar to other ADM studies^9,18,23^, we first performed separate single-ancestry analyses in two admixed sub cohorts within AoU: individuals with genetically predicted Admixed American (n=40,327; 17.5% of AoU) and African (n=49,797; 21.6% of AoU) ancestries. We also repeated the ADM in a multi-ancestry framework by pooling all participants into a single analysis (n = 230,019; **Fig. 2a**) which we describe below. For the Admixed American cohort, we performed association testing for both NAT and AFR local ancestry labels, given the representation of both ancestries in that group. For the African cohort, which is largely admixed between African and European ancestries, we performed association testing for AFR local ancestry only. In each case, we tested for association between the number of haplotype copies assigned to that label in each individual (0, 1, or 2) in each window and the trait. Notably, we did not perform separate tests for EUR local ancestry, since the number of EUR segments in a window is highly anticorrelated with the number of AFR or NAT segments making the EUR tests largely redundant, and the EUR label showed high discrepancy across tools (**Fig. S1**).

**Figure 2:**
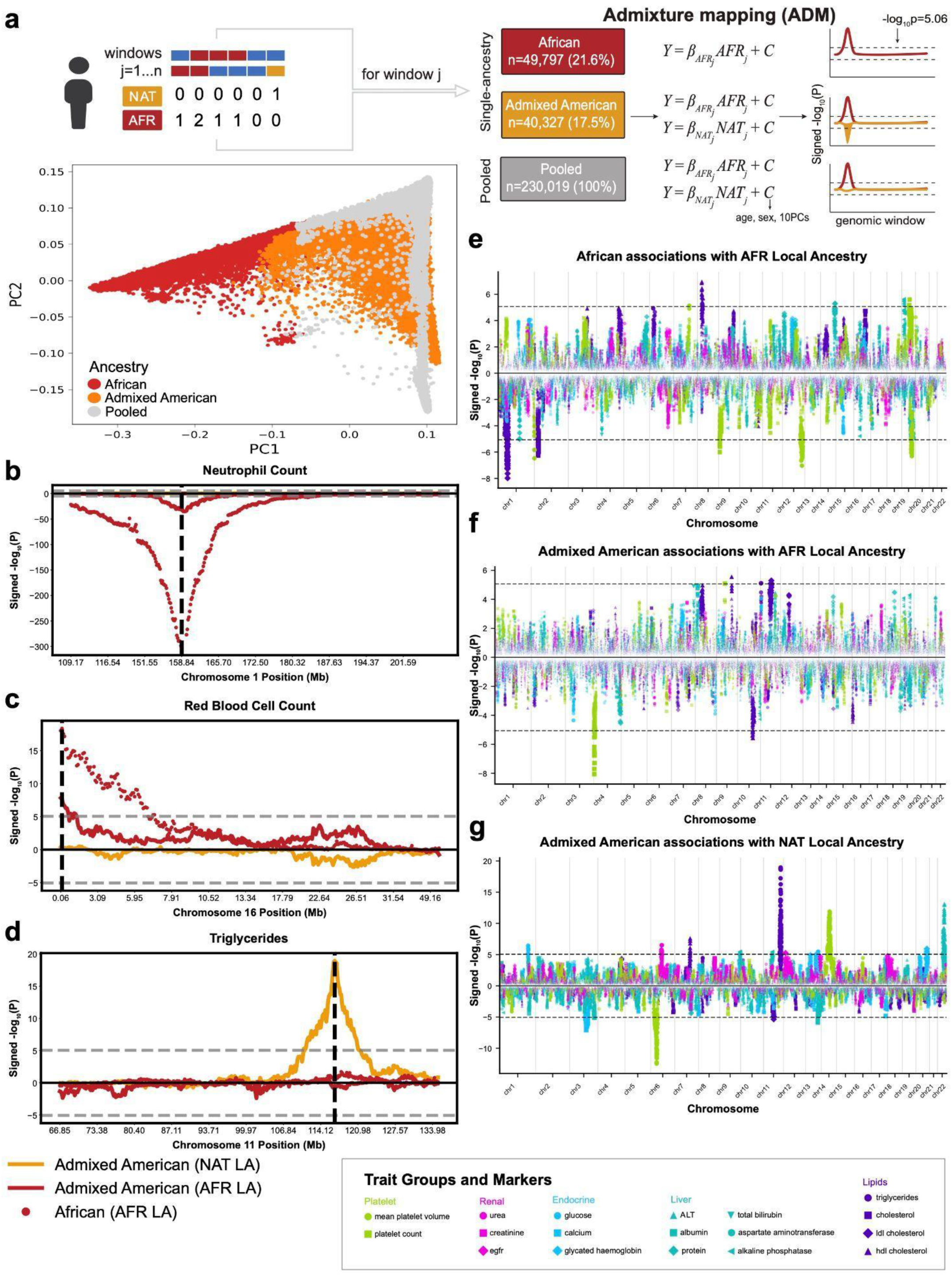
Admixture mapping (ADM) workflow and sample sizes in the *All of Us Research Program*. (a) PCA plot and schematic of the admixture mapping workflow. For each genomic window, we tested for association between the number of copies of local ancestry (NAT or AFR) and each trait within single ancestry cohorts and the pooled cohort (comprising all genotyped multi-ancestry AoU samples). The genetically predicted African cohort consisted of 49,797 individuals, and the Admixed American cohort included 40,327 individuals. The pooled multi-ancestry analysis included all 230,019 samples. Sample sizes for association testing varied across the 29 traits depending on phenotypic data availability and QC (**Table S2**). **(b-d) Known admixture mapping signals captured by our pipeline.** We recovered known associations of AFR local ancestry at the *ACKR1* (Duffy) locus associated with neutrophil count **(b)**, AFR local ancestry at the α-globin locus associated with red blood cell count **(c)**, and NAT local ancestry at the *BUD13/APOE5/ZPR1* locus associated with triglycerides **(d)**. Dashed horizontal lines denote the genome-wide -log10 P-value significance threshold of 5.06. The *y*-axis shows the -log10 P-value with the signed effect direction for each local ancestry label tested (red=AFR, gold=NAT). **(e-g) Summary of ADM results for single ancestry cohorts.** Panels **e-g** show ADM results based on the African cohort for AFR local ancestry **(e)**, the Admixed American for AFR local ancestry **(f)** and the Admixed American for NAT local ancestry **(g)**. In each panel the *x*-axis gives the genomic coordinate, and the *y*-axis gives the −log₁₀(P-value) with the signed effect direction for each GNOMIX window. Dot colors denote the trait category (grass green=platelet, pink=renal, blue=endocrine, aqua blue=liver, purple=lipids) and shape denotes individual traits. Red blood cell and white blood cell count results are shown separately (**Fig. S3-S4**) due to the extremely strong signals for those traits.

We next established a genome-wide significance threshold for the ADM association tests performed here. Notably, because local ancestry labels can have extensive long-range linkage disequilibrium (LD)^24^, a simple Bonferroni correction for the number of bins tested is overly conservative. Thus, we implemented a permutation approach to empirically determine an appropriate threshold, adopting a previously used strategy^25^ (**Methods**). Notably, this approach is highly compute-intensive and thus could not be applied to each trait tested. Instead, we applied this approach on two representative quantitative traits for multiple settings (including single-ancestry and pooled, see below and **Methods**). Across all configurations tested, the significance thresholds (corresponding to an empirical P < 0.05) ranged from a -log10 P-value of 4.91 to 5.13 (**Table S3**). Based on these results, we established a study-wide significance threshold of -log10 P-value = 5.06 (P=8.7 × 10⁻⁶) corresponding to the median threshold across configurations. Note, this is still considerably stricter than thresholds used in similar admixture mapping analyses (e.g. -log10 P-value = 4.3 in Mandla *et al.*^18^).

In the single ancestry analyses, we identified a total of 53 independent trait-region associations in the Admixed American cohort (32 for NAT, 21 for AFR) and 44 associations in the African cohort meeting genome-wide significance and passing all quality filters (**Table S4; Methods**). Top signals in both groups capture multiple known local ancestry associations (**Table 1**; **Fig. 2b-d**), including associations between AFR local ancestry at the *ACKR1* (Duffy) locus on Chromosome 1 with neutrophil count^9^ (P<5.65e−36 in Admixed Americans, P<1e-300 in Africans; **Fig. 2b**) and other white blood cell traits; between AFR local ancestry at the α-globin locus on Chromosome 16 with red blood cell count^26^ (P=1.47e−8 in Admixed Americans, P=4.22e-19 in Africans, **Fig. 2c**); between NAT local ancestry at a locus on Chromosome 11 overlapping *BUD13/APOE5/ZPR1*^23^ with triglycerides (P=1.12e−19 in Admixed Americans; **Fig. 2d**); and between NAT local ancestry at a locus on Chromosome 14 overlapping *ACTN1*^27^ with mean platelet volume (P<1.30e−12 in Admixed Americans). All but 5 signals overlapped with genes reported in the NHGRI GWAS Catalog to be associated with these or similar traits (**Table S5**), although most of these signals have not previously been identified to be associated with local ancestry (e.g. see examples in **Table 1**).

**Table 1.** Top admixture mapping association signals in the African and Admixed American cohorts. The table summarizes loci with the most significant associations identified for AFR local ancestry (top 5 rows) and NAT local ancestry (bottom 5 rows). For each locus, the table lists the genomic coordinates (GRCh38) of the lead GNOMIX bin, genes overlapping the lead bin, associated traits, the discovery cohort (African and/or Admixed American), the signed-log10 P-value for each significantly associated trait, the local ancestry label (AFR or NAT), and whether the signal has been identified by previous admixture mapping studies. For some regions where the lead GNOMIX bin is near but not overlapping known signals, the second column notes the name of the known gene or region in parentheses.

| Table 1: Top local ancestry associations |  |  |  |  |  |  |
| --- | --- | --- | --- | --- | --- | --- |
| AFR associations |  |  |  |  |  |  |
| Lead region (hg38) | Overlapping genes | Trait(s) | Cohort(s) | local ancestry associated (AFR/NAT) | signed $-\log_{10}$ (P) (African, Admixed American) | Known ADM signal? |
| chr1:159154758-159328573 | *ACKR1* | WBC<br>Neutrophil ct.<br>Lymphocyte %<br>Neutrophil %<br>Eosinophil % | African, Admixed Americans | AFR | -300, -72.7 (WBC)<br>-301, -35.2 (Neutrophil ct.)<br>274, 41.4 (Lymphocyte %)<br>-229.1, -36.9 (Neutrophil %)<br>42.34, 6.38 (Eosinophil %) | Yes<sup>9</sup> |
| chr16:10164-126890 | *POLR3K*,<br>*SNRNP25*,<br>*RHBDF1*, *MPG*,<br>*NPRL3* (alpha globin locus) | RBC<br>RBCDW<br>MCV<br>MCH<br>Hemoglobin | African, Admixed Americans | AFR | 18.37, 7.83 (RBC)<br>12.14, 8.12 (RBCDW)<br>-55.69, -35.92 (MCV)<br>-66.86, -43.53 (MCH)<br>-5.91, -5.70 (Hemoglobin) | Yes<sup>30</sup> |
| chr4:3274232-3439943 | *RGS12* | Platelet count | Admixed American | AFR | -8.06 | No |
| chr1:57031138-57179245 | *DAB1 (near PCSK9 locus)* | LDL cholesterol<br>cholesterol | African | AFR | -7.99 (LDL cholesterol)<br>-6.59 (cholesterol) | Yes<sup>31</sup> |
| chr2:158738882-158872526 | *DAPL1* | WBC | African | AFR | 7.05 | No |
| **NAT associations** |  |  |  |  |  |  |
| chr11:116840367-117025856 | *SIK3, (BUD13, APOE5, ZPR1 locus)* | Triglycerides<br>cholesterol | Admixed American | NAT | 18.95 (triglycerides)<br>5.63 (cholesterol) | Yes<sup>23</sup> |
| chr22:43891707-43976310 | *PNPLA3, SAMM50, PNPLA5* | ALT<br>AST | Admixed American | NAT | 13.14 (ALT)<br>6.45 (AST) | Yes<sup>32</sup> |
| chr6:36526101-36682957 | *STK38, CDKN1A, SRSF3 (near HLA)* | MPV | Admixed American | NAT | -12.47 | No |
| chr14:67163086-67334315 | *GPHN, GARIN2, PALS1, ATP6V1D (ACTN1 locus)* | MPV | Admixed American | NAT | 11.88 | Yes<sup>33</sup> |
| chr7:101296746-101437997 | *COL26A1, IFT22* | HDL<br>cholesterol | Admixed American | NAT | 7.60 | No |

In many instances, we found that the same genomic locus was associated with multiple distinct genes and traits. Combined across the Admixed American and African cohorts, there were 65 total AFR local ancestry-driven signals, which mapped to 20 unique genomic regions. Similarly, 32 significant associations driven by NAT ancestry corresponded to 17 distinct genomic loci. Many of these shared associations highlight the correlated genetic architecture of these traits. For NAT local ancestry, the *PNPLA3* locus was associated with liver-related traits, specifically AST, ALT, and glycated hemoglobin (which has been previously linked to liver disease^28,29^). For AFR local ancestry, the *ACKR1* (Duffy) and α-globin loci were strongly associated with multiple white and red blood cell traits, respectively (**Fig. S3–S4**). A notable exception is the HLA region, where local ancestry in the Admixed American population was significantly associated with two distinct traits: AFR local ancestry with mean platelet volume, and NAT local ancestry with mean corpuscular volume.

### Comparison of AFR local ancestry associations in Admixed American vs. African cohorts

We next evaluated the consistency of AFR local ancestry-driven effects across the Admixed American and African cohorts, which consist of completely separate individuals and therefore serve both as replication cohorts but also enable exploring potential cohort-specific signals. We considered all 65 independent signals passing genome-wide significance in at least one of the two cohorts. Overall, 52.3% of signals identified in the African cohort showed at least nominal (P<0.05) significance in the Admixed American cohort. Similarly, 71.4% of signals identified in the Admixed American cohort were at least nominally significant in the African cohort (**Fig. 3a**). Signals identified in both groups tended to have overall stronger P-values than those identified in only a single cohort (**Fig. 3b**). We then compared effect sizes across cohorts. All 65 signals showed concordant directions of effects across the two cohorts (**Fig. 3c-e**). The magnitude of effect sizes for white blood cell and red blood cell traits were highly concordant across cohorts (Pearson r=0.98 and 0.92 for WBC and RBC respectively), which is primarily driven by the extremely strong effects of the *ACKR1* (Duffy) and alpha globin loci. Association statistics for all other trait categories showed more modest but significant correlation (Pearson r=0.59, two-sided P=0.02; **Fig. 3e**). We attribute this to the statistical lower power to detect weaker signals particularly in the Admixed American cohort which have overall lower percentages of AFR local ancestry in all 65 bins (**Table S6**), consistent with the genome-wide average AFR local ancestry across the two groups. Despite the lower correlation of effect sizes, effect directions were concordant across cohorts for 100% of the loci tested. Overall, our results demonstrate the robustness of these signals across cohorts and suggest the majority of AFR local ancestry signals are shared across African and Admixed Americans.

**Figure 3.**
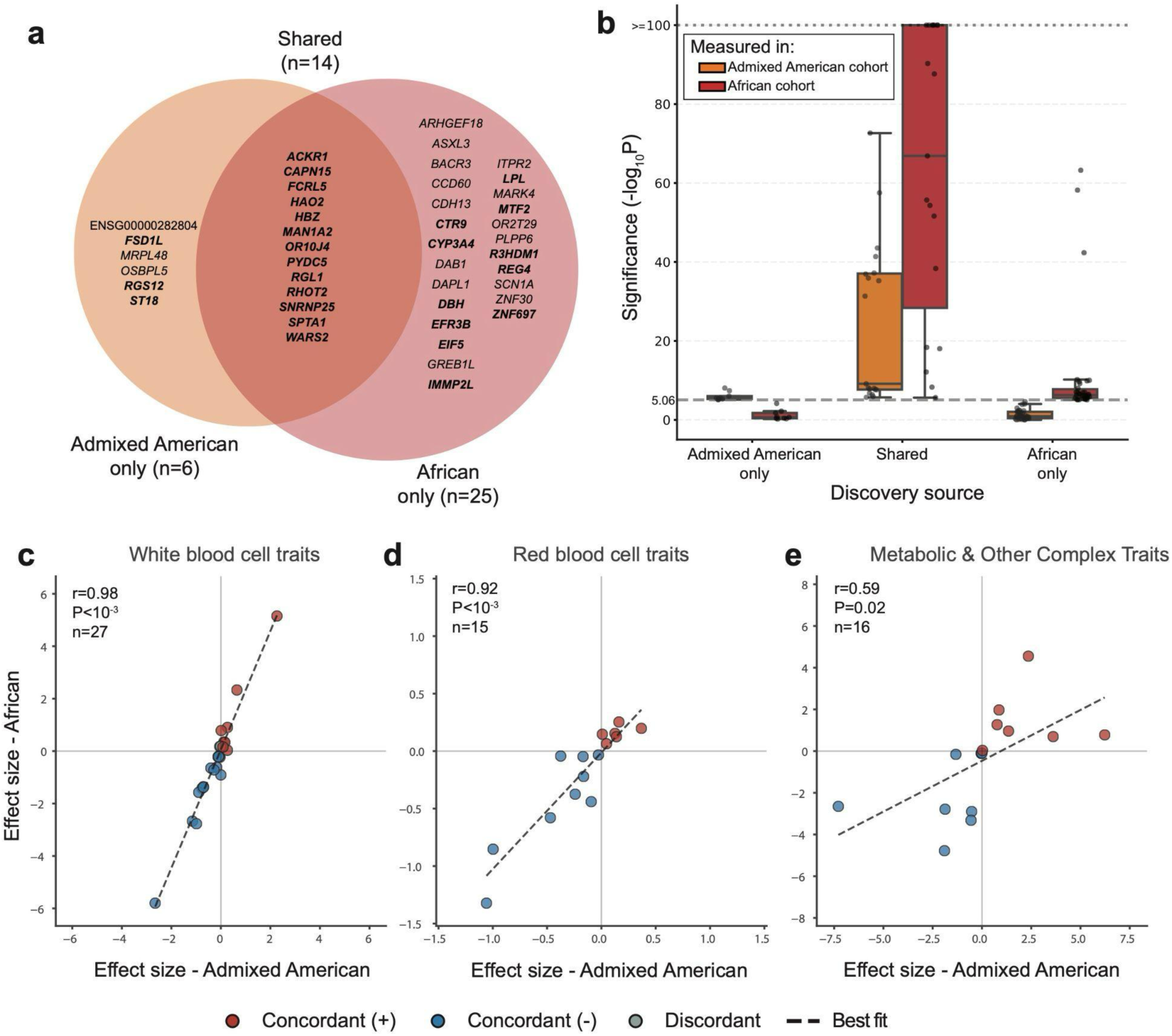
Evaluating the robustness of AFR local ancestry-trait associations in Admixed American and African cohorts. (a). Venn diagram illustrating the intersection of trait-region associations with AFR local ancestry. The golden circle represents associations identified at genome-wide significance only in the Admixed American cohort (*n* = 6), while the red circle represents associations identified only in the African cohort (*n* = 25 unique loci). The orange intersection (*n* = 14) denotes loci with significant effects detected in both cohorts. Associations are annotated by their nearest gene. Bolded genes indicate loci that reached nominal significance in the other group. E.g. bolded loci in the golden circle only passed genome-wide significance in Admixed Americans but were nominally significant in Africans. **(b). Distribution of association statistics -log10 P-value stratified by discovery source.** Distributions are shown for trait-region associations identified either in Admixed American only (left), both cohorts (middle), or Africans only (right). Golden=Admixed American P-values, red=African P-values. The dashed horizontal line indicates the genome-wide significance threshold (-log10 P-value =5.06). Dotted horizontal line indicates -log10 P-value=100. Values were capped at 100 for display purposes. **(c-e). Comparison of effect sizes between the Admixed American (x-axis) and African (y-axis) cohorts for AFR local ancestry associations.** Only trait-region associations passing genome-wide significance in at least one of the two cohorts are included. Plots are stratified by trait category: **(c)** white blood cell traits, **(d)** red blood cell traits, and **(e)** metabolic & other complex traits. Points are colored by directional concordance: red=concordant positive effects; blue=concordant negative effects; grey=effects with discordant direction. Note, all signals showed concordant directions, so no grey dots are included. Pearson correlation, two-sided *P*-values, and the number of data points (n) are annotated in each plot.

Although most of our signals were concordant across cohorts—with generally more significant effects in Africans—we observed several AFR local ancestry signals that showed substantially stronger effects in Admixed Americans. These include unknown signal on Chromosome 4 for platelet count overlapping *RGS12* (P = 8.76×10⁻⁹ in Admixed American and P = 0.00607 in African; **Fig. S5**) and a signal on Chromosome 11 for triglycerides overlapping the *OSBPL5/MRGPRG/MRGPRE* region (P = 7.83×10⁻⁶ in Admixed American and P = 0.400 in African; **Fig. S6**) which has been previously implicated in cardiometabolic traits in Hispanic/Latino populations^34^. Both signals could not be explained by differences in the frequency of AFR local ancestry at these windows, which were consistent with genome-wide averages (**Table S6**). These findings highlight the complexity of cross-population replication, even when focusing on shared local ancestry labels. In addition, they suggest that although a signal may be driven by AFR local ancestry, that signal might show different effects across cohorts with different global ancestries.

### Analysis of ancestry-specific enrichment of ADM signals across trait categories

To understand the broader relationship between the 97 ADM signals and the ancestral origin of complex traits, we aggregated related phenotypes into seven major biological categories—endocrine, renal, liver, lipid, white blood cell (WBC), red blood cell (RBC), and platelet traits—and quantified the distribution of signals by local ancestry (**Fig. 4**). This analysis revealed a distinct divergence in signals attributed to NAT vs. AFR local ancestry across trait types. Metabolic and organ function traits tended to show associations with NAT local ancestry; specifically, endocrine traits showed a near-significant enrichment of NAT signals (5/5; P = 0.062, two-sided binomial test), and liver and renal traits showed a similar directional bias (5/7; P = 0.453 for liver and 2/2; P=0.50 for renal). In contrast, the majority of associations identified for hematologic traits were with AFR local ancestry. This enrichment was almost statistically significant for white blood cell traits (11/14; P = 0.057) and showed consistent biases but did not reach significance for red blood cell (9/15; P = 0.136) and platelet (6/8; P=0.289) traits. Lipid traits displayed a more balanced pattern, with signals attributable to both ancestries (8 AFR, 4 NAT, P = 0.388). Some of these differences correspond to overall differences in the distribution of phenotype across cohorts. For example, ALT and glucose have significantly higher values in the Admixed American vs. African cohorts (Mann-Whitney two-sided P=1.2e-153 and P=5.3e-46 respectively, **Fig. S2**) and tend to be associated with NAT local ancestry. On the other hand, platelet count, which has only AFR local ancestry signals, shows similar distributions across cohorts (P=0.65).

**Figure 4:**
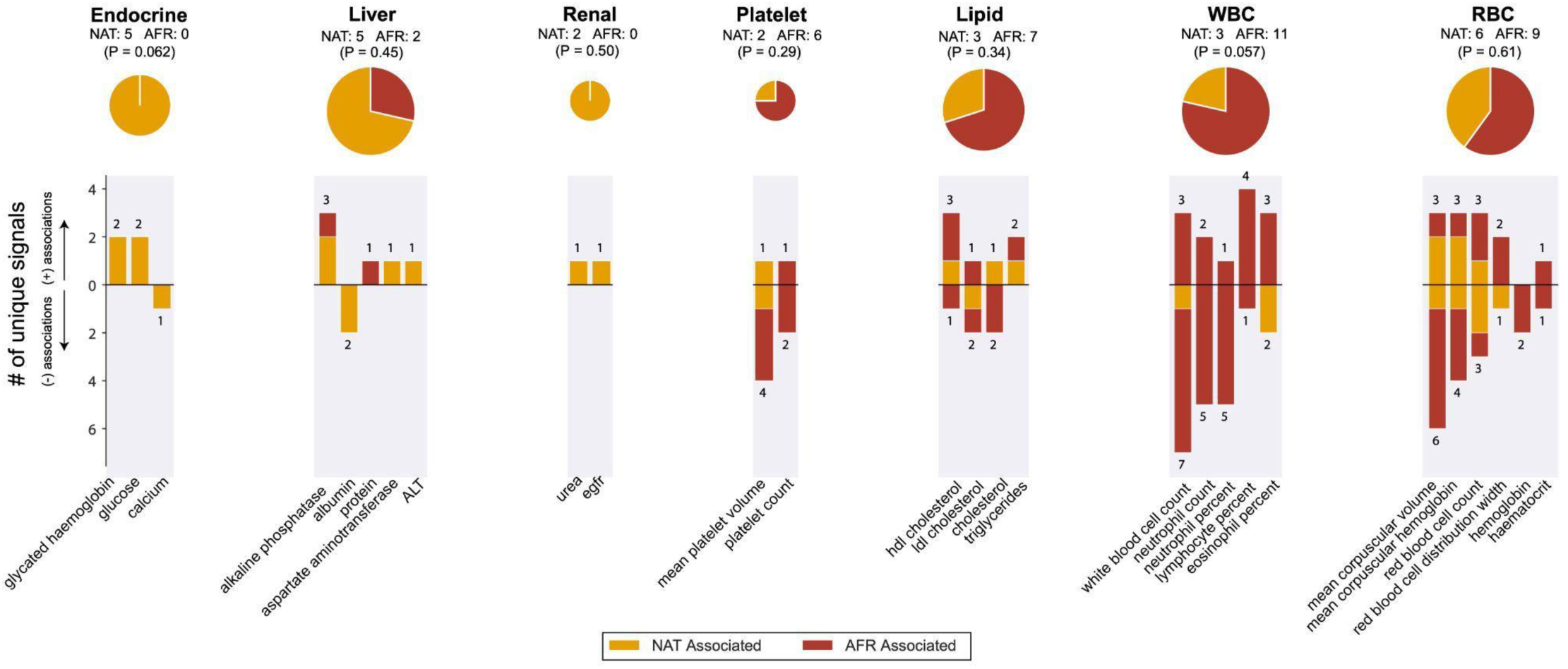
Distribution of ADM signals by local ancestry and trait category. Each panel summarizes the number of unique genome-wide significant ADM associations per trait obtained in the single ancestry association analysis (Admixed American and African cohorts), grouped into broader biological categories: endocrine, liver, renal, platelet, lipid, white blood cell (WBC) and red blood cell (RBC) traits. Within each panel, individual bars represent specific traits, with bar height indicating the number of associations and the effect direction of the bar (upward or downward) reflecting the sign of the effect size (positive or negative association, respectively, of the indicated local ancestry with the trait). Associations are color-coded by ancestry (red=AFR, gold=NAT). Pie charts above each panel show the total proportion of AFR versus NAT local ancestry associations across all traits within the corresponding category. P-values are based on a two-sided binomial test. For the trait-category summaries (pie charts and P-values), loci that showed associations across multiple traits were collapsed to a single data point to avoid counting potentially pleiotropic effects from the same locus (e.g. Duffy with WBC traits) multiple times.

Within each category, we further examined the direction of effect for each signal. For endocrine trait signals, signals associated with NAT local ancestry tended to be associated with elevated glucose and glycated hemoglobin, as well as lower calcium levels, all of which have been associated with higher risk for metabolic diseases such as type 2 diabetes^35–37^. On the other hand, significant AFR signals for lipids tended to show associations between AFR local ancestry in those regions with elevated HDL and lower LDL, both of which are associated with protective effects for cardiovascular disease^38^. Overall, these findings highlight substantial differences in traits associated with NAT vs. AFR local ancestry and highlight how local ancestry may shape disease risk differently across present-day admixed populations (**Discussion**).

### Admixture mapping across the entire AoU cohort

Finally, we extended ADM to a multi-ancestry context by jointly analyzing all participants (n = 230,019). We performed separate association testing in this cohort for NAT and AFR local ancestry, again encoding each individual as having 0, 1, or 2 copies of each label in each bin. Compared to the analyses presented above, which consider only 39% of available participants, the pooled strategy allows including 100% of participants (**Fig. 2a**) regardless of predicted global ancestry. This approach has the potential to improve power due to larger sample sizes, but may be prone to confounding by population structure or other factors^39,40^ which we explore below.

This combined analysis identified 261 independent region-trait associations meeting genome-wide significance (P = 8.7 × 10⁻⁶) for either AFR (n=135 signals) or NAT (n=127 signals) local ancestry (**Fig. S8**, **Table S4**). Consistent with the single-ancestry analyses (**Fig. 2b-d**), known loci for WBC and RBC traits remained among the top signals (**Fig. S3–S4**). Similar numbers of pleiotropic loci were observed, with 28 independent regions associated with more than one trait in NAT and AFR, respectively. Finally, we observed a similar trend in which signals for metabolic and organ function tended to be associated with NAT local ancestry whereas blood cell trait signals tended to be driven by AFR local ancestry (**Fig. S7**).

Performing ADM in a pooled setting identified 2.7-fold more hits than in the single ancestry analyses presented above (261 vs. 97). We reasoned that the increased number of signals could be driven by a combination of increased power due to larger sample size as well as false positive signals due to uncorrected population stratification in a highly admixed cohort. To investigate this further, we quantified the inflation of test statistics (-log10 P-values) across both the single-ancestry and pooled results for each trait using the genomic inflation factor (*λ_GC_*; **Methods; Table S7; Fig. 5a-b**). Indeed, while the inflation was modest for many traits, we observed across all settings that test statistics were more at least modestly more inflated in the pooled setting. Notably, *λ_GC_* varied widely across traits suggesting that susceptibility to population stratification is not uniform but rather depends on the specific trait and ancestry combinations studied. We observed that association tests for AFR local ancestry showed less overall inflation with several notable exceptions, primarily related to hematological traits. For example, the difference in *λ_GC_* across the pooled vs. single-ancestry setting (termed Δλ) was 1.03 for mean corpuscular hemoglobin and 0.90 for platelet count. In contrast, we observed the most severe inflation for associations with NAT local ancestry, particularly in metabolic and organ function traits; Δλ was 2.16 for glucose and 1.87 for glycated hemoglobin, with similar differences observed for renal and liver function traits. Overall, these observations suggest that the genetic architecture or environmental confounders related to certain traits are uniquely stratified within the African vs. Admixed American populations.

**Figure 5:**
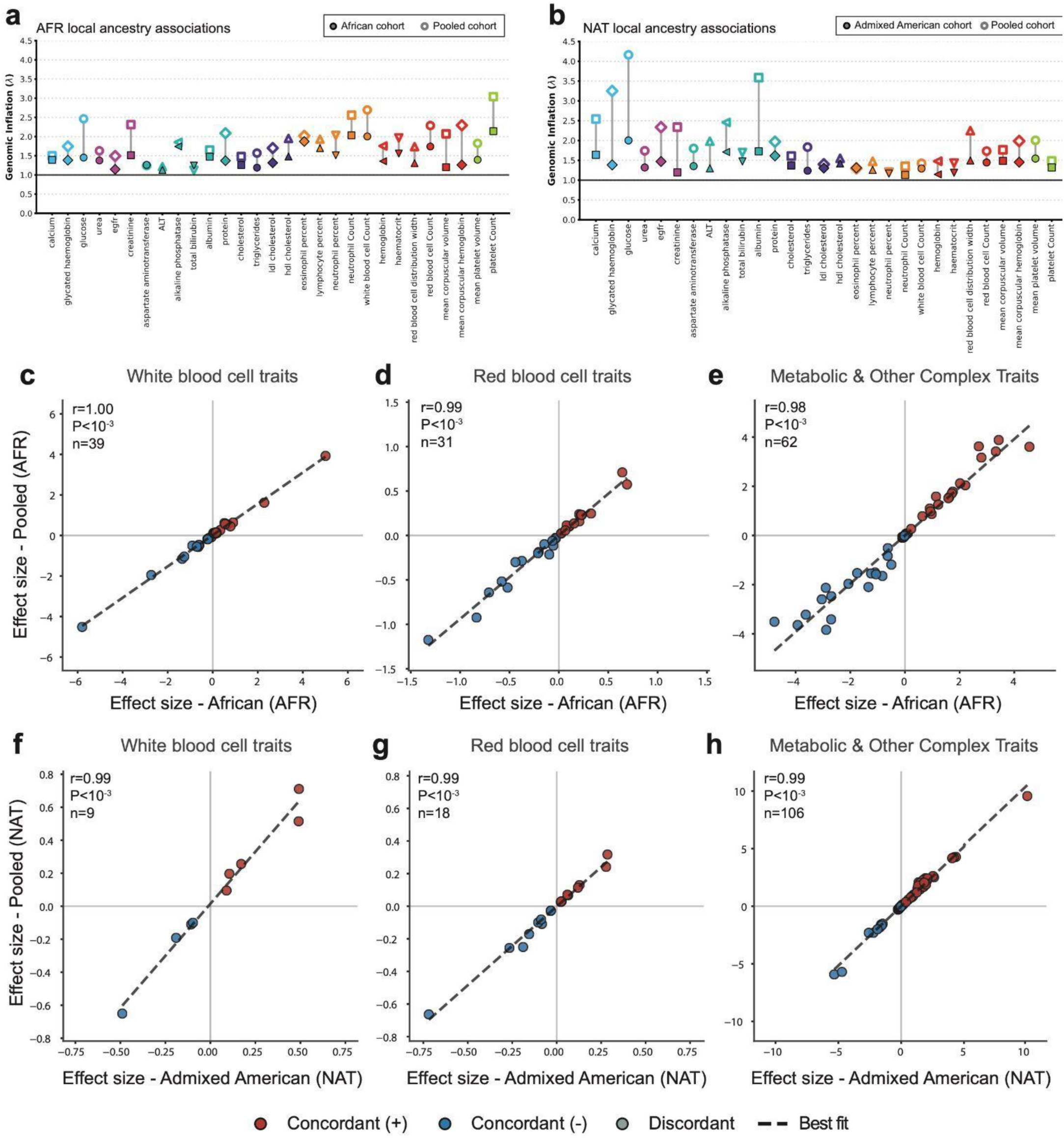
Comparison of ADM results in pooled vs. single-ancestry cohorts. (a-b) Comparison of genomic inflation (λ) across analyzed traits organized by trait groups. **(a)** shows results for AFR association testing in the African (filled shapes) vs. pooled (open shapes) cohorts. **(b)** shows results for NAT association testing in the Admixed American (filled shapes) vs. pooled (open shapes) cohorts. The black horizontal line at y=1 indicates no inflation. (**c-h) Comparison of effect sizes in single ancestry (x-axis) vs. pooled (y-axis) cohorts.** Only trait-region associations passing genome-wide significance in at least one of the two cohorts compared are included. Plots are stratified by trait category: **(c, f)** white blood cell traits, **(d, g)** red blood cell traits, and **(e, h)** metabolic & other complex traits. **(c-e)** compare AFR local ancestry signals in the African vs. pooled cohorts and **(f-h)** compare NAT local ancestry signals in the Admixed American vs. pooled cohorts. Points are colored by directional concordance: red=concordant positive effects; blue=concordant negative effects; grey=effects with discordant direction. Note, all signals showed concordant directions, so no grey dots are included. Pearson correlation, two-sided *P*-values, and the number of data points (n) are annotated in each plot.

To further evaluate the reliability of signals observed in the pooled analysis, we directly compared summary statistics for trait-region associations identified at genome-wide significance in either the pooled or single-ancestry analyses (**Fig. 5c-h; Fig. S9**). Effect sizes showed remarkable correlation across settings (Pearson r > 0.98 across all traits and local ancestry labels considered), with 100% of associations showing concordant direction of effect. Effect sizes showed similar magnitude across pooled vs. ancestry-specific analysis. Similarly, -log10 P-values were highly consistent (r>0.92 in all cases; **Fig. S9**) with a tendency for slightly stronger P-values in the pooled analysis. The strong correlation of effect sizes as well as the reproducibility of top hits and overall trends observed in the single-ancestry analyses suggest that although P-values from the pooled analysis are subject to inflation – likely due to the introduction of additional uncorrected population structure (**Discussion**) – the rank order of associations is robust, and signals are highly reproducible across analysis settings. Overall, our results suggest the pooled analysis, which enables inclusion of all individuals regardless of predicted ancestry, is a powerful strategy for performing analysis in multi-ancestry cohorts, but likely requires additional techniques to adjust for confounding to ensure additional signals identified are true vs. driven by uncorrected population structure.

## Discussion

This study represents, to our knowledge, the largest admixture mapping study to date. We analyzed individuals from the *All of Us* Research Program with genetically predicted Admixed American (n=49,797) and African (n=40,327) ancestries, as well as a pooled cohort (n=230,019) consisting of the entire AoU v7 cohort. Using GNOMIX to infer local ancestry, we tested 29 clinically relevant quantitative traits for association with African (AFR) and Native American (NAT) ancestry segments. In the single-ancestry analysis, we identified 97 significant local ancestry–trait associations spanning 37 distinct genomic loci. Several associations recapitulate well-established findings, including African ancestry in the *DARC* (Duffy antigen)^9^ region associated with white blood cell counts and related phenotypes, and NAT local ancestry at the *BUD13/APOE/ZPR1* locus associated with triglyceride levels^23^, whereas others were not previously identified (**Table 1**). Identified signals are highly robust across cohorts, with highly correlated effect sizes (**Fig. 3c-e**; **Fig. 5c-h**) and 100% concordance of effect direction of signals for the same local ancestry label in separate cohorts (AFR in African vs. Admixed Americans) and for signals identified in single-ancestry vs. a pooled setting.

The majority of signals identified here by admixture mapping overlap genes previously associated with similar traits through variant-level GWAS (**Table S5**). We hypothesize that the majority of ADM signals are driven by individual causal variants with large differences in allele frequencies across populations. The causal variants likely have causal effects across all populations^41^, but are detected as local ancestry-specific signals for the ancestry background on which they are most common. In these cases, single-variant tests as performed in GWAS should also capture the signal and be better powered than ADM unless local ancestry is perfectly linked with the causal variant. However, such signals still are often missed by standard GWAS pipelines for several reasons. GWAS performed in relatively homogeneous ancestry cohorts may exclude or miss ancestry-specific variants due to low minor allele frequencies. For example, the lead variant at the Duffy locus has a MAF<0.5% in Europeans. Even when included power to detect the association will be lower in the populations where the variant is less common. An additional challenge is that ancestry specific variants tend to show strong departures from expected genotype frequencies under Hardy Weinberg Equilibrium, especially when considering multi-ancestry cohorts^42^, and thus are often not considered.

On the other hand, 5.2% of signals identified by our ADM approach did not overlap known GWAS signals. These could be explained by a variety of scenarios. One possibility is that the signal is driven by an ancestry-specific variant that was not directly genotyped and is better tagged by local ancestry than by any individual genotyped variant. One such example is at the well-known *LPA* locus associated with lipoprotein(a) concentration [Lp(a)].

Previous work found that local ancestry is strongly associated with Lp(a) and that common SNPs in the region show associations with Lp(a) in European but not African populations^43^. However, this is likely driven by the fact that the main causal variant in the region is a highly polymorphic kringle(IV) tandem repeat which shows highly divergent repeat length distributions across populations^44^ and is differentially tagged by bi-allelic SNPs in Africans vs. Europeans^43^. Another potential explanation is that some local ancestry signals may be driven by interactions between variants in *cis* on haplotypes enriched in certain ancestries. A third possibility is that local ancestry signals are driven by interactions with non-genetic factors or with other genomic regions. Identifying the specific causal factors underlying these signals is challenging and represents an important area of future work.

Our comparison of AFR local ancestry associations in African and Admixed American individuals revealed high consistency across these groups. All effects that reached significance in at least one cohort showed concordant direction of effect across both cohorts, and effect sizes were highly correlated across all trait categories (**Fig. 3c-e**). We observed stronger P-values in the African cohort, which we hypothesize is largely driven by reduced power in the Admixed American cohort due to the lower overall frequency of AFR local ancestry, rather than biological differences. However, we also identified two notable exceptions for which AFR local ancestry signals were much stronger in Admixed Americans, specifically at the *RGS12* locus for platelet count (**Fig. S5**) and the *OSBPL5/MRGPRG* region for triglycerides (**Fig. S6**) which has been previously implicated in cardiometabolic traits in Hispanic/Latino populations^34^. These findings could not be explained by differences in AFR local ancestry frequency at those loci. One potential explanation for this discrepancy is that the AFR haplotypes in the Admixed American cohort at this locus carry distinct genetic variants from AFR haplotypes in the African. Alternative explanations include potential interaction with other ancestry-specific variants in *trans*, or parent of origin effects at the *OSBPL5* locus, which overlaps the chr11p15.5-15.4 region known to exhibit imprinting^45^. Overall, these results suggest that while most AFR local ancestry effects are consistent across different admixed cohorts, specific signals might be influenced by the different genetic backgrounds or environments unique to each population. Further work is needed to determine the specific mechanism leading to the ancestry-specific effects observed here.

When grouping traits into broader biological categories, we observed strong patterns of enrichment for signals associated with NAT vs. AFR local ancestry across trait groups. Signals for hematological traits were disproportionately associated with AFR local ancestry at specific genomic regions. These patterns may reflect selection associated with environmental exposures across populations. For example, the Duffy-null allele provides resistance to malaria, and has risen to high frequency in African populations but is not prevalent in other populations where malaria is uncommon^46^. Further, signals in the HLA locus may be associated with historical exposures of different populations to other pathogens over time^47^. On the other hand, signals for endocrine, renal, and liver traits were disproportionately associated with NAT local ancestry at specific genomic regions. These findings are consistent with the elevated burden of type 2 diabetes^48^, hypertension^49^, and chronic kidney disease^50^ observed in Hispanic/Latino populations. While these disparities are likely driven by a combination of genetic and non-genetic factors^51^ such as diet, lifestyle, or exposure to pathogens, our results suggest that the higher rates of these conditions in certain populations may be driven in part by ancestry-specific genetic variants within the ADM signals identified here.

Finally, we evaluated the feasibility of performing admixture mapping in the entire AoU v7 multi-ancestry cohort, rather than restricting to individual admixed groups (Admixed Americans and Africans) as has been done in previous studies. Compared to the single-ancestry approach, the pooled strategy has multiple benefits including improving power, not requiring excluding individuals based on predicted ancestry, and simplifying analysis since all local ancestry labels can be tested in the same cohort. On the other hand, this strategy may be subject to false positives due to uncorrected population structure. A recent study comparing these approaches for GWAS in AoU and other cohorts demonstrated that the pooled strategy generally exhibited higher power and tests were generally well calibrated, but that test statistics could be subject to inflation if a substantial fraction of phenotypic variation is explained by within-ancestry principal components^52^. We observed similar trends here: the pooled analyses resulted in nearly three times as many signals as the single-ancestry analyses, with signals showing high concordance with but stronger P-values compared to those obtained from single-ancestry analyses (**Fig. 5c-e, Fig. S7-9**), but in some cases showing inflated test statistics. While the inflation was modest for many traits, it was particularly severe for NAT local ancestry associations with metabolic traits like glucose and renal function. This suggests that combining multi-ancestry groups introduces population structure for some traits that is not fully accounted for by genotype principal components. Taken together, our results demonstrate that while pooling individuals is a powerful strategy for discovery, it requires careful attention to population structure to ensure that the additional signals represent true genetic associations; furthermore, they highlight the need for methodological improvements such as spectral components^39^ or other approaches, to better correct for confounding factors, particularly given the increasing admixture and population structure within evolving multi-ancestry biobanks which cannot be fully captured by the standard approach of using genetic principal components.

Our study faced several limitations. First, although the AoU cohort contains individuals from a wide range of ancestries, our ADM study was limited to testing the most prevalent non-European local ancestry labels (AFR and NAT). We excluded testing for associations with East Asian or South Asian local ancestry due to low sample size. Additionally, we did not test for associations with West Asian ancestry due to a combination of low sample size and highly discordant results for that label across tools (**Fig. S1**). Second, the local ancestry inference approaches used here require a set of reference haplotypes that ultimately relies on discrete population labels. Both global and local ancestry represent continuous properties that cannot be fully described by these labels. Further, the existing high-level labels remove information about substructure and haplotype diversity within each group. Third, our study only focused on 29 quantitative traits. Additional insights are likely to be gained by expanding our ADM pipeline in future work to additional disease phenotypes.

Overall, our study demonstrates how admixed populations offer a unique opportunity to discover signals missed by standard approaches and by restricting to relatively homogeneous populations. The United States is a profoundly admixed nation and the number of individuals who identify as admixed continues to grow^53^. As larger multi-ancestry cohorts become available, analysis approaches that fully leverage admixture are likely to discover additional signals and improve the utility of downstream precision genomics applications such as risk prediction across ancestry groups.

## Methods

### Sample and variant data preprocessing

We performed analysis of samples from all available ancestries in the *All of Us* version 7.1 release using the short-read whole genome sequences (srWGS) v7.1 ACAF dataset consisting of 99,250,816 SNPs and indels (at 48,314,438 sites) with allele frequency greater than 1% or allele count greater than 100 in any predicted ancestry subpopulation. We excluded samples with call rates below 90%. For each sample, we also computed the transition/transversion ratio, heterozygous/homozygous ratio, insertion/deletion ratio, and number of singletons and filtered samples whose value for any of these metrics falls outside 8 standard deviations of the mean across all samples. We excluded 15,375 related samples flagged by *All of Us* using pre-computed kinship scores by Hail (relatedness_flagged_samples.tsv) to keep a maximum set of unrelated participants. We used samples that self-report “male” or “female” for sex assigned at birth. Self-reported race, ethnicity and predicted ancestry classifications for each sample were obtained from the file ancestry_preds.tsv provided by All of Us. Finally, admixture mapping was performed on all remaining samples. We additionally provide phenotype summary statistics for various ancestry cohorts for comparison in **Table S2 and Fig. S2**. The African and Admixed American cohorts are based on predicted ancestry (individuals with “afr” or “amr” in the column “ancestry_pred” in ancestry_preds.tsv). The pooled cohort consists of all individuals passing quality control steps above regardless of predicted ancestry.

### Local ancestry inference with GNOMIX

We applied GNOMIX^16^ using their published pretrained models downloaded from GitHub (**URLs**) for local ancestry inference. We focused on the available model rather than building a custom model, which is a compute-intensive process and for which we encountered memory constraints that could not be resolved. The pretrained model includes the following superpopulations: Subsaharan African (AFR), East Asian (EAS), European (EUR), Native American (NAT), Oceanian (OCE), South Asian (SAS), West Asian (WAS) and Ancestral Hunter Gatherers (AHG).

Prior to local ancestry inference, to avoid excessive memory use with downstream tools we first created separate VCF files for each chromosome for each batch of 1000 samples. We then used bcftools v1.20^54^ with the liftover plugin^55^ to convert VCF files for each batch from GRCh38 to hg19 coordinates because available GNOMIX models are only available for this reference build. We used Beagle version 5.4^56^ to perform phasing and imputation (option impute=true) using the 1000 Genomes Phase 3 reference panel^19^. The resulting phased imputed VCF files were further split to batches of 100 samples each using the bcftools split plugin. We applied GNOMIX^16^ to each batch of samples using their pretrained models described above., with the phase option set to False. Finally, we used a custom script to concatenate Gnomix results for all samples for each chromosome.

### Local ancestry inference with FLARE

To run FLARE, we used a merged VCF for 245,394 samples from the srWGS ACAF dataset described above and similarly removed related samples leaving 230,019 unrelated samples. Multi-allelic variants were split into biallelic variants using bcftools v1.20, resulting in one record per alternate allele. We performed reference-free phasing using BEAGLE^56^ v5.4 with window size 10-20 cM varied by chromosome length due to memory limitations in the AoU environment. We parallelized each run per chromosome across five separate AoU Cloud workspaces. We used phased VCF files as input to local ancestry calling with FLARE v0.5.2. We combined both HGDP and 1KGP to create a reference panel of 2,265 samples, which includes six ancestry groups: European (GBR, FIN, IBS, CEU, TSI; n=525), Non-Admixed African (GWD, ESN, MSL, LWK, YRI; n=529), South Asian (PJL, BEB, STU, ITU, GIH; n=512), and East Asian (CHS, CDX, KHV, CHB, JPT; n=512) in 1000 Genomes, plus West Asian (Druze, Bedouin, Palestinian; n=133), and Native American (Surui, Maya, Karitiana, Pima; n=54) from the HGDP to ensure that each population consists of homogeneous samples similar to related studies^17,57,58^. To improve accuracy suggested by the original developers of FLARE, we created population-specific VCF files using global ancestry prediction values precomputed with a random forest classifier based on principal components provided by AoU. To manage computational loads, any subset containing more than 10,000 samples was further partitioned into multiple chunks of approximately 10,000 individuals each. For instance, on a typical chromosome, EAS (n=5,525 samples) required only one chunk, whereas EUR (n=128,302 samples) was split into 14 chunks. We ran FLARE separately on each chunked VCF. Once FLARE finished running on all chunks for a given chromosome, we merged those chunk-level output files to produce unified files that include all 230,019 unrelated AoU samples.

### Global ancestry inference

We used the same reference panel as constructed above for FLARE for global ancestry inference using genotype files obtained from the plink2 resources webpage (**URLs**). For global ancestry analysis, we used plink2^59^ to subset the srWGS ACAF dataset described above to filter variants with call rate below 5%, minor allele frequency below 5%, and those deviating from Hardy-Weinberg equilibrium (P<1×10^-6^). Similar to suggestions in the ADMIXTURE manual and Rye manuscript, we then used the plink2 indep-pairwise function to select independent SNPs using a window size of 50kb, step size of 2 variants, and an LD r^2^ threshold of 0.2 and harmonized pruned variants from AoU and the reference panel files. Overall, we were able to retain 83,922 shared variants after the SNPs harmonization of the AoU and the reference panel for downstream analysis.

We used two tools for global ancestry inference: Rye^22^ v0.1 and ADMIXTURE^21^ v1.3. For Rye, we used plink2 --pca to compute the eigenvectors and eigenvalues jointly on the harmonized reference and AoU samples. We retrieved the first 10 eigenvectors and used these as input to Rye with non-default options pcs = 10, round = 100, and inter = 100. For ADMIXTURE, we first used the reference panel .bed file from plink2 as input with option --supervised to calculate variant allele frequencies across the six reference populations. We used these as input to project the AoU samples onto the reference using the -P option to ADMIXTURE.

To infer global ancestry percentages from GNOMIX, for each individual we summed the length of all windows assigned to each local ancestry label and divided this by the total length of all windows (considering both chromosome copies on Chromosomes 1-22). To infer global ancestry from FLARE, we used its chromosome-level ancestry proportion outputs and computed a genome-wide, chromosome length-weighted average for each ancestry per individual.

### Phenotype preprocessing

The All of Us Research Program leveraged the Observational Medical Outcomes Partnership (OMOP) common data model v5 for a harmonized phenotype data representation of electronic health record (EHR) data submitted by multiple institutions^60^. OMOP concept IDs for each of the 29 quantitative traits analyzed are shown in **Table S2.** For each trait, after restricting to data points from individuals passing sample preprocessing above and matching one of the units listed in **Table S2**, we filtered values more than 5 standard deviations away from the mean. We then obtained the median phenotype value per year that measurements were available for each sample, then took the median of medians across all years, leaving at one most data point per individual per phenotype. We included the age at the time of measurements as a covariate in the regression model for association testing. From the standardize serum creatinine (*S_cr_*) value, we calculated estimated glomerular filtration rate (eGFR) using CKD-EPI formula 3^61,62^: 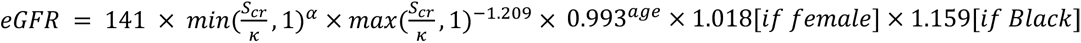, where *S_cr_* is serum creatinine level in mg/dL; *κ* is 0.7 for females and 0.9 for males; and *α* is -0.429 for females and -0.411 for males.

### Admixture mapping in the All of Us Research Program

We performed admixture mapping to identify associations between NAT and AFR local ancestry and each of the 29 biomarkers on the genetically predicted Admixed American, African cohort, and a multi-ancestry pooled cohort. For each GNOMIX window we encoded ancestry (AFR or NAT) dosage as 2 if an individual had two copies of the ancestry at that window, 1 for one copy, and 0 if the ancestry was not present. We then used a linear regression model to test each ancestry separately for association with the trait, adjusting for age, sex, and the first 10 genotype principal components as covariates (**Fig. 2a**).

We used a permutation approach to determine an empirical genome-wide significance threshold. Prior to the permutation analysis, we filtered out GNOMIX windows that failed liftover from GRCh38 to hg19 coordinates or resulted in genomic regions spanning more than 250kb as these regions could be prone to mapping artifacts. We performed permutation analysis on two different traits (RBC, ALT) across all 5 ADM configurations considered (AFR in Pooled, African, and Admixed American cohorts and NAT in the Admixed American cohort). In each case, we permuted phenotype values and repeated ADM. We repeated this procedure 1,000 times, in each case recording the top P-value. We then determined an empirical -log10 P-value threshold as that corresponding to the 95th percentile across all 1,000 simulations in each setting. Since we obtained similar results across settings, we chose the median across all runs, corresponding to -log10P=5.06, as the threshold throughout the manuscript. For each trait, for each ancestry label tested in each cohort, we used a clumping procedure to determine the number of independent significant signals. For this, we applied the following steps: (1) take the window with the strongest P-value; (2) extend the region to window_start-25Mb through window_end+25Mb, add all windows in that region to the current clump and remove from further consideration; (3) repeat steps 1-2 until no windows passing the significance threshold remain. For downstream analyses, resulting significant windows were re-converted to GRCh38 coordinates using the same liftOver plugin (**URLs**)^55^. Following coordinate conversion to GRCh38, gene annotations were assigned using GENCODE Release 44 (**URLs**) to identify gene features overlapping the significant genomic windows (**Table S5**).

### Overlapping ADM signals with the NHGRI-EBI GWAS Catalog

To evaluate whether the genes overlapping identified candidate loci from ADM had been previously associated with similar traits based on genome-wide association studies (GWAS), we queried the full NHGRI-EBI GWAS Catalog (**URLs**). The dataset was programmatically retrieved from the EBI FTP server on November 25, 2025. We extracted the ’MAPPED_GENE’ and ‘DISEASE/TRAIT’ columns and used these to construct a list of traits associated with each gene. For each ADM signal, we determined the list of genes overlapping the lead GNOMIX bin. **Table S5** shows associated traits from the GWAS Catalog for each gene in each window.

## URLs

GNOMIX pretrained models: https://github.com/AI-sandbox/gnomix/blob/main/download_pretrained_models.sh plink2 resources for references samples: https://www.cog-genomics.org/plink/2.0/resources

Rye V1.0: https://github.com/healthdisparities/rye

ADMIXTURE V1.3: https://dalexander.github.io/admixture/download.html

UCSC liftOver plugin: http://hgdownload.cse.ucsc.edu/goldenpath/hg19/liftOver/hg19ToHg38.over.chain.gz GENCODE v44 (GRCh38):

https://ftp.ebi.ac.uk/pub/databases/gencode/Gencode_human/release_44/gencode.v44.annotation.gtf.gz NHGRI-EBI GWAS Catalog: ftp.ebi.ac.uk/pub/databases/gwas/releases/latest/

## Data availability

This study used data from the *All of Us Research Program*’s Controlled Tier Dataset V7.1, available to authorized users on the Research Workbench.

## Code availability

Workflows for this project are available on GitHub: https://github.com/cast-genomics/cast-workflows.

## Author contributions

W.G.G.R led, designed and performed LAI with GNOMIX, ADM on single and pooled analysis and wrote the manuscript. Y.L. performed phasing with BEAGLE and LAI with FLARE. T. M. developed pipelines for quality filtering in AoU samples. N. M. performed phenotype preprocessing and helped with pipeline setup. J.K helped supervise BEAGLE and FLARE analyses. M.A. and U.D. helped with the manuscript. K.A.F. helped supervise the study and manuscript. M.G. helped develop pipelines for GNOMIX LAI, supervised the study and wrote the manuscript.

## Acknowledgements

This research was supported in part by the NIH/NHGRI grant RM1HG011558 (M.G. and K.A.F.). W.G.G.R was supported by the Alfred P. Sloan Foundation Program [G-2022-10127], the ARCS foundation, and NIH grant T15LM011271. We thank Rahel Wachs for her illustrations that supported the communications of our findings. We gratefully acknowledge *All of Us* participants for their contributions, without whom this research would not have been possible. We also thank the National Institutes of Health’s *All of Us Research Program* for making available the participant data examined in this study.

