## Supplementary Figures for "Admixture mapping identifies complex trait associations with local ancestry in the *All of Us Research Program*"

#### Figure S1: Pairwise comparison of estimated global ancestry percentages inferred by ADMIXTURE, Rye, GNOMIX, and FLARE.

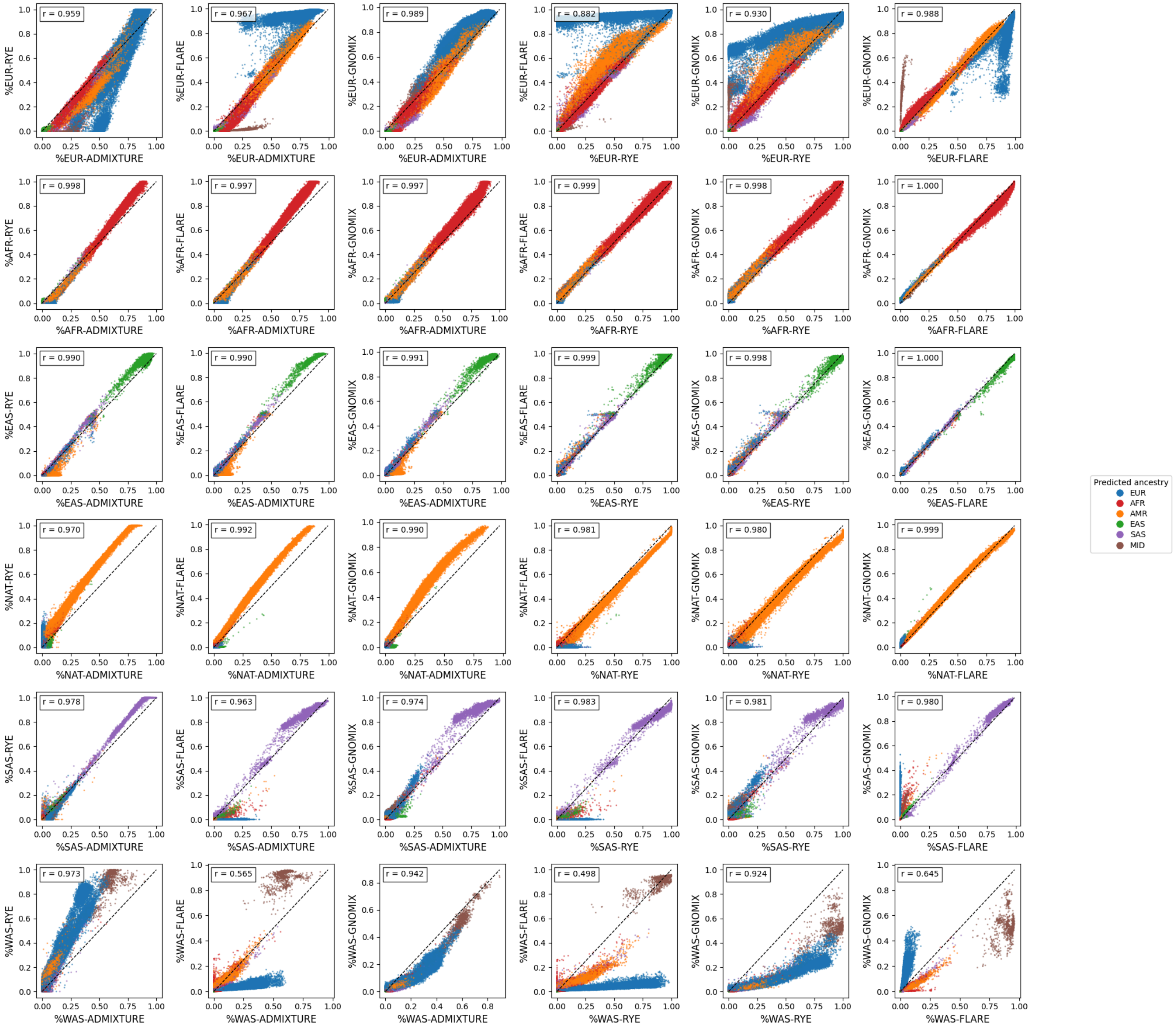


Plots show inferred global ancestry percentages based on all four tools for each individual, colored by predicted ancestry labels provided by All of Us. Black lines give the x=y diagonal.

#### Figure S2: Phenotype distributions in the Admixed American and African cohorts

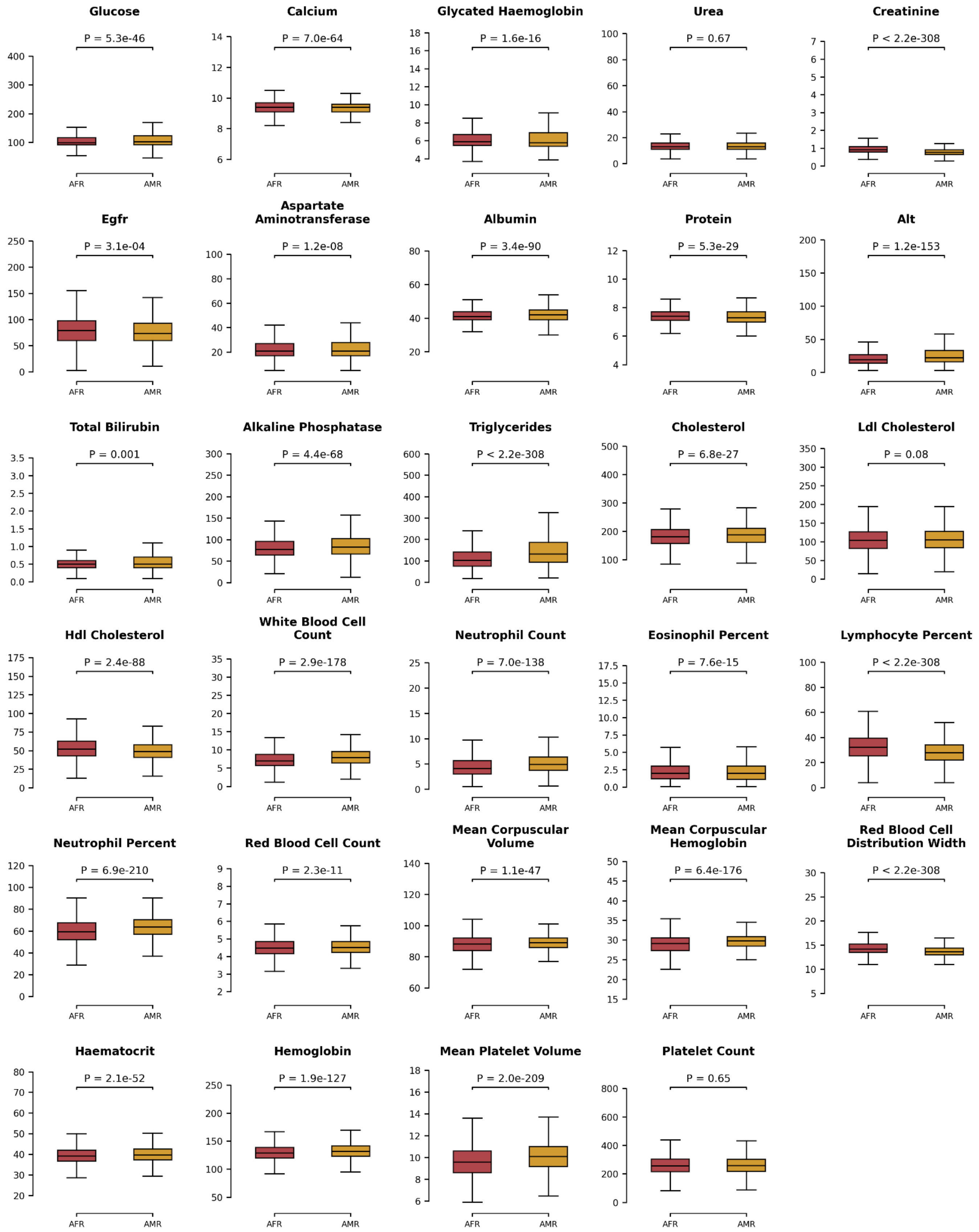


Boxplots illustrate the distribution of phenotype values for individuals in the African (AFR) and Admixed American (AMR) ancestry cohorts. The center line represents the median, box limits indicate the upper and lower quartiles (25th and 75th percentiles), and whiskers extend to 1.5× the interquartile range (IQR). Statistical significance was assessed using a two-sided Mann-Whitney *U* test.

#### Figure S3: Whole genome admixture mapping results for WBC traits

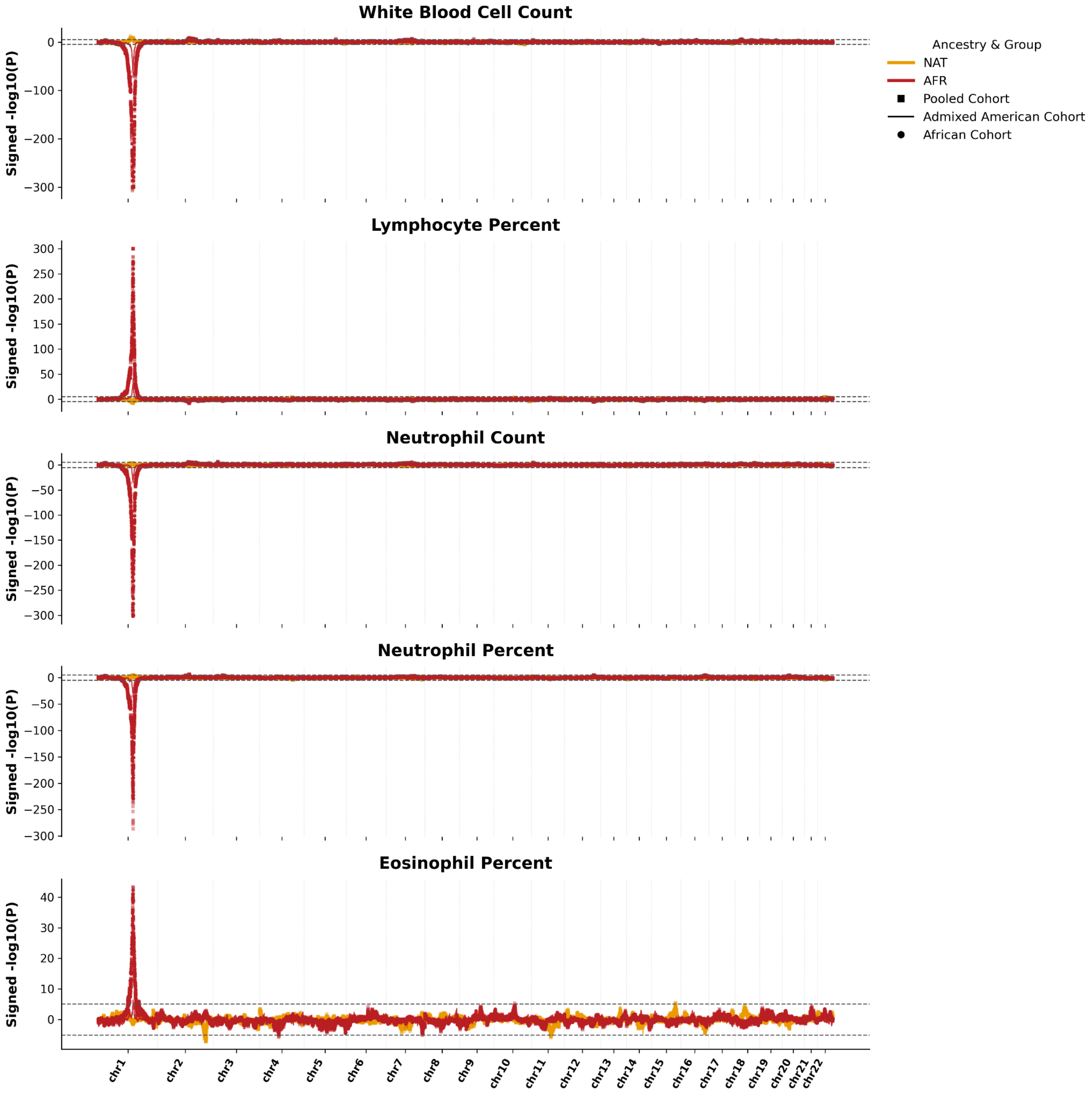


Panels show admixture mapping results for WBC-related traits for all cohorts. In each panel the *x*-axis gives the genomic coordinate and the *y*-axis gives the signed -log10 P-value for each region based on predefined windows provided by GNOMIX for each ancestry label tested (red=AFR, gold=NAT). Dashed horizontal lines denote the -log10 P-value significance threshold of 5.06. Squares=pooled cohort; circles=African cohort; lines=Admixed American cohort.

#### Figure S4: Whole genome admixture mapping results for RBC traits.

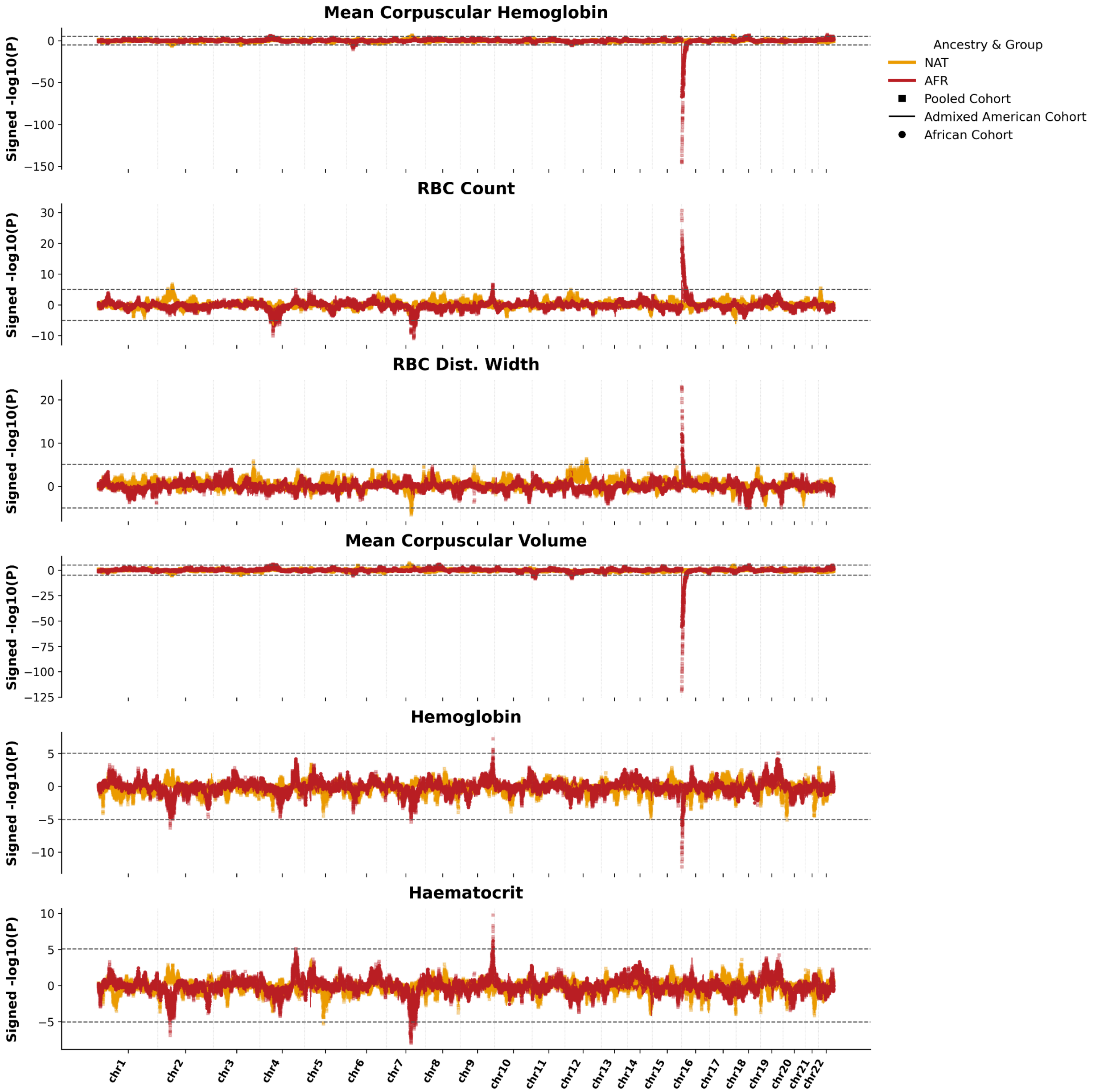


Panels show admixture mapping results for RBC-related traits for all cohorts. In each panel the *x*-axis gives the genomic coordinate and the *y*-axis gives the signed -log10 P-value for each region based on predefined windows provided by GNOMIX for each ancestry label tested (red=AFR, gold=NAT). Dashed horizontal lines denote the -log10 P-value significance threshold of 5.06. Squares=pooled cohort; circles=African cohort; lines=Admixed American cohort.

**Figure S5: Local ancestry association signal for Platelet Count on Chromosome 4**

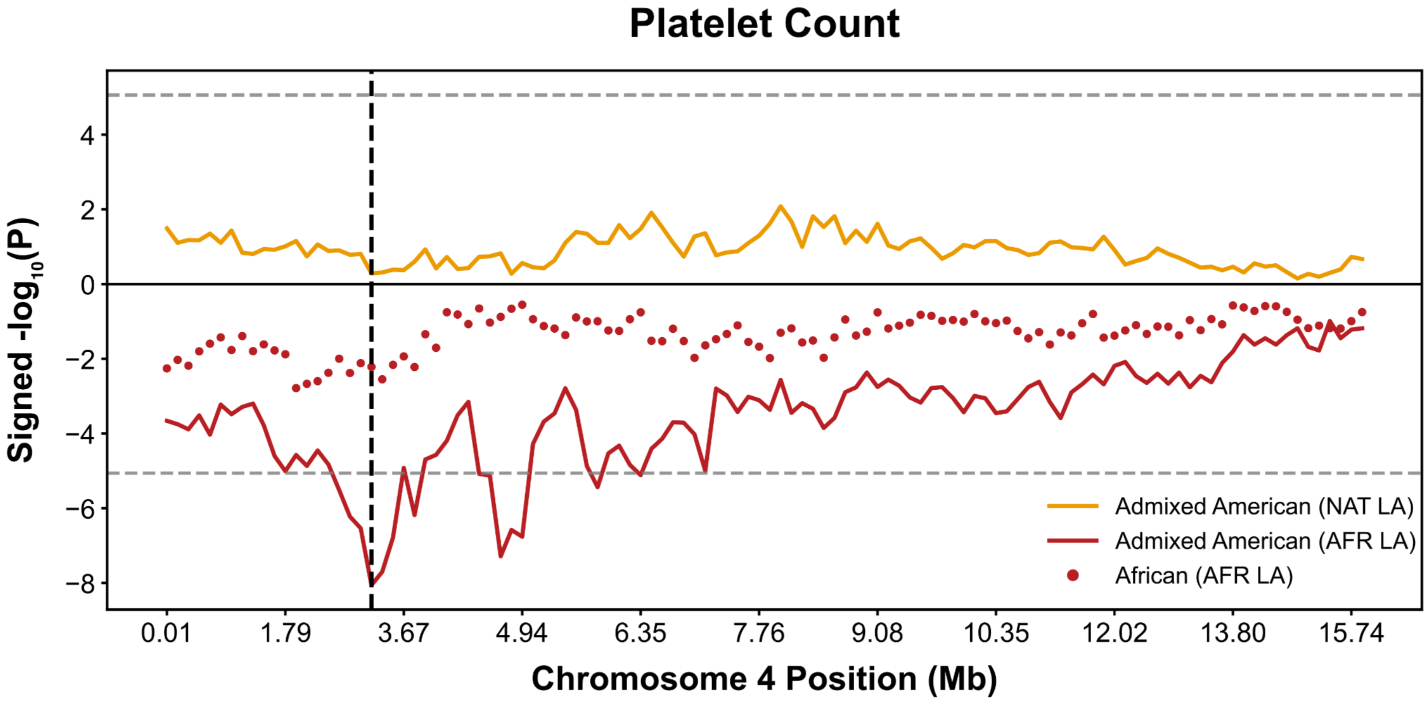


Admixture mapping plot displaying the local ancestry association signal for platelet count overlapping the gene *RGS12* on Chromosome 4. The *y*-axis shows the signed -log10 P-value for each local ancestry label tested (red=AFR, gold=NAT). The vertical black dashed line marks the position of the lead bin. Horizontal gray dashed lines indicate the genome-wide significance threshold. Lines=associations in the Admixed American cohort; circles=associations in the African cohort.

#### Figure S6: Local ancestry association signal for Triglycerides on Chromosome 11

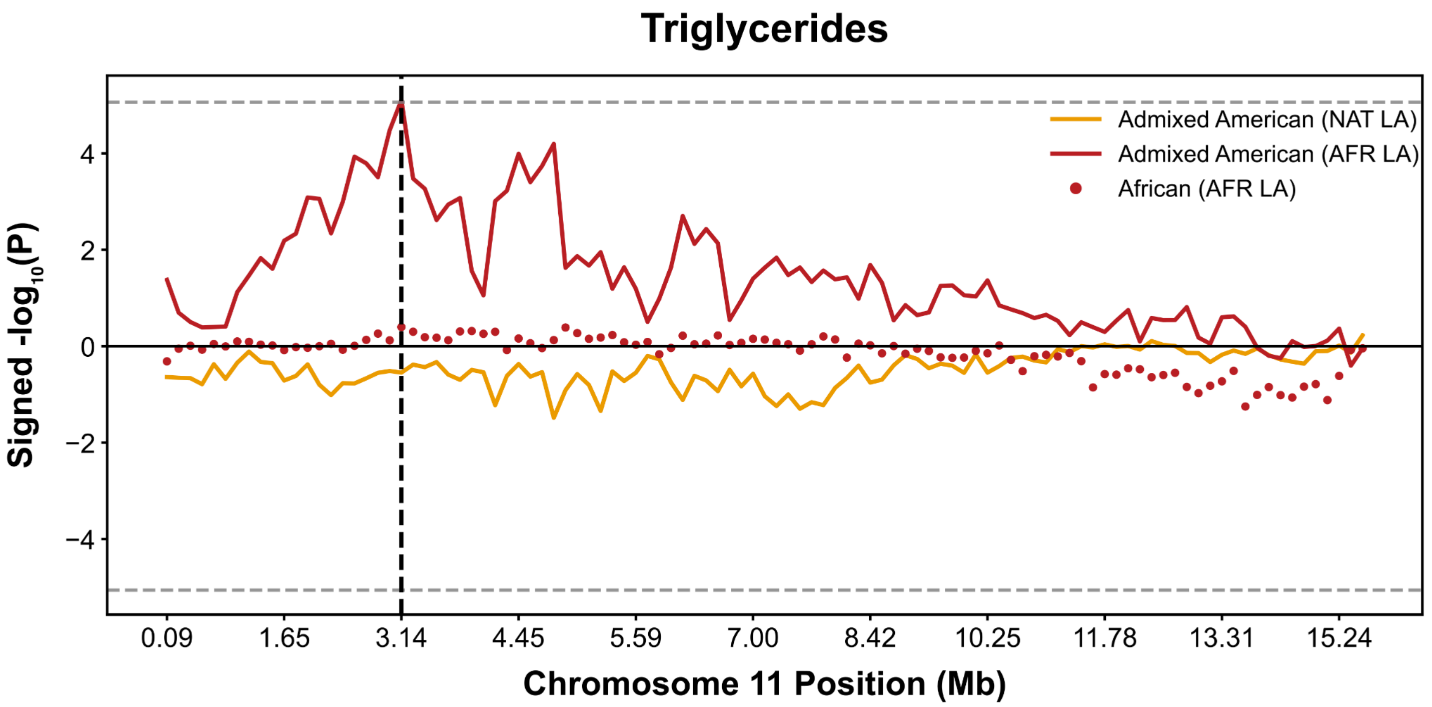


Admixture mapping plot displaying the local ancestry association signal for triglycerides at region overlapping genes *OSBPL5*/*MRGPRG/MRGPRE* chromosome 11. The *y*-axis shows the signed -log10 P-value for each local ancestry label tested (red=AFR, gold=NAT). The vertical black dashed line marks the position of the lead bin. Horizontal gray dashed lines indicate the genome-wide significance threshold. Lines=associations in the Admixed American cohort; circles=associations in the African cohort.

#### Figure S7: Distribution of admixture mapping signals by local ancestry and trait category in the pooled cohort

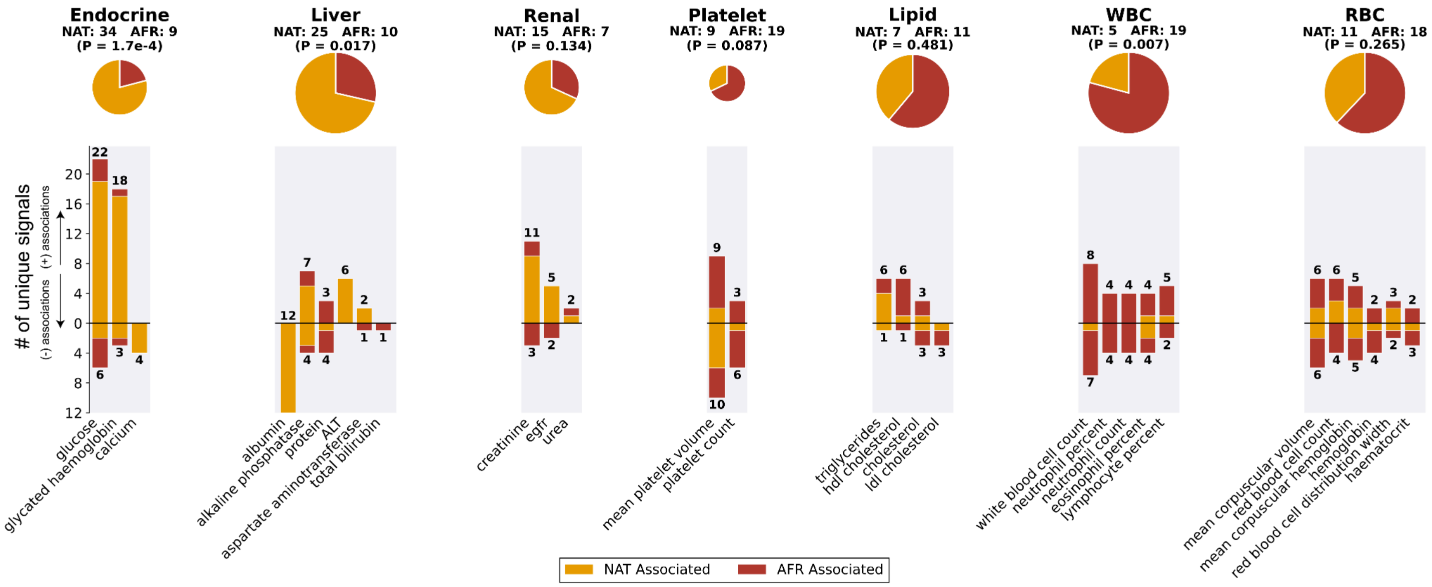


Each panel summarizes the number of unique genome-wide significant ADM associations per trait obtained in the pooled ancestry association analysis grouped into broader biological categories: endocrine, liver, renal, platelet, lipid, white blood cell (WBC) and red blood cell (RBC) traits. Within each panel, individual bars represent specific traits, with bar height indicating the number of associations and the effect direction of the bar (upward or downward) reflecting the sign of the effect size (positive or negative association, respectively, of the indicated local ancestry with the trait). Associations are color-coded by ancestry (red=AFR, gold=NAT). Pie charts above each panel show the total proportion of AFR versus NAT local ancestry associations across all traits within the corresponding category. P-values are based on a two-sided binomial test. For the trait-category summaries (pie charts and P-values), loci that showed associations across multiple traits were collapsed to a single data point to avoid counting potentially pleiotropic effects from the same locus (e.g. Duffy with WBC traits) multiple times.

#### Figure S8: Summary of ADM results in cohort-specific and pooled analyses

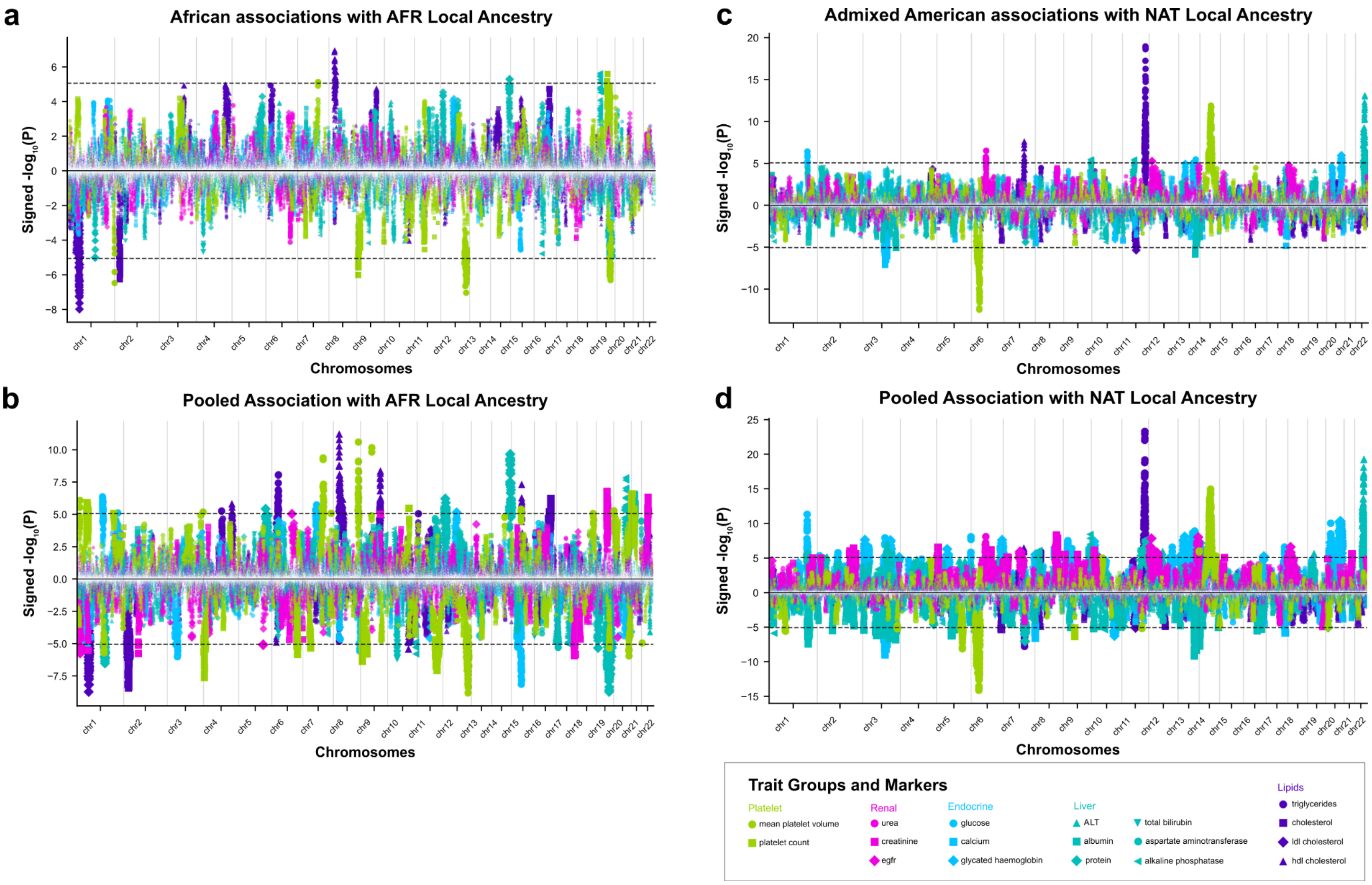


Panels show admixture mapping associations for the African cohort with AFR local ancestry (**a**), the pooled cohort with AFR local ancestry (**b**), the Admixed American cohort with NAT local ancestry (**c**), and the pooled cohort with NAT local ancestry (**d**). In each panel, the x-axis gives the genomic coordinate and the y-axis gives the signed -log10 P-value for each GNOMIX window. Dot colors denote the trait category (green = platelet, pink = renal, light blue = endocrine, teal = liver, purple = lipids) and shape denotes individual traits. Dashed horizontal lines indicate the genome-wide significance threshold. Red blood cell and white blood cell count results are shown separately (**Fig. S3-S4**) due to the extremely strong signals for those traits.

#### Figure S9: Consistency of association signals between pooled and cohort-specific analyses

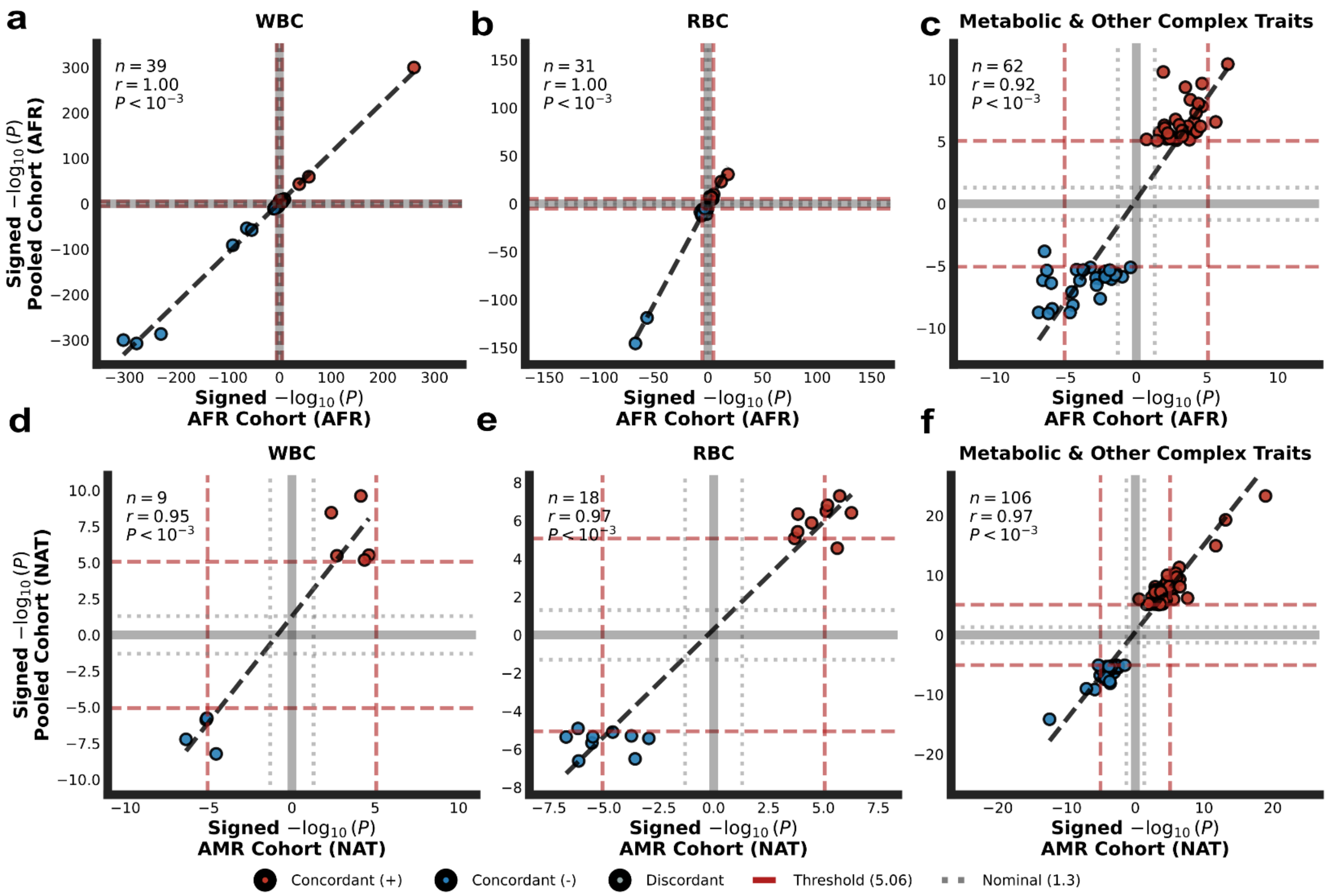


Comparison of signed -log10 P-value values between the pooled analysis (y-axis) and single ancestry cohort analyses (x-axis) across trait categories: left=White Blood Cell (WBC) count, middle=Red Blood Cell (RBC) count, and right=Metabolic & Other Complex Traits. **Top panels (a–c)** Comparisons of the AFR local ancestry signals between the pooled analysis and the African (AFR) cohort. **Bottom panels (d–f)** Comparisons of the NAT local ancestry component signals between the pooled analysis and the Admixed American (AMR) cohort. Points are colored by concordance: red indicates concordant positive effects, and blue indicates concordant negative effects. The dashed diagonal line represents the line of identity (x=y), indicating perfect replication of signal magnitude and direction. Grey dotted lines represent the nominal significance threshold (P = 0.05). Red dashed lines denote the genome-wide significance threshold. The numbers of data points (n), Pearson correlation coefficients (r) and corresponding two-sided P-values are displayed in the upper-left corner of each panel.

### SUPPLEMENTARY TABLE LEGENDS

#### Table S1: Estimated global ancestry proportions based on self-reported race/ethnicity

This table shows the estimated global ancestry proportions for each self-reported race or ethnicity for each tool. Column headers are in the format <ancestry>_<tool>_glob.

#### Table S2: Summary of the traits used for admixture mapping

This table provides descriptive statistics for all traits included in the admixture mapping analysis. For each trait, the table reports the trait category, concept ID, units, cohort, cohort size, sex distribution (male and female ratios), mean participant age, mean phenotype value, phenotype standard deviation, and minimum and maximum observed values. For comparison, we also show phenotype summary statistics for different ancestry cohorts in **Fig. S2**.

#### Table S3: Empirical significance thresholds determined by permutation testing

Summary of empirical genome-wide significance thresholds (-log10 P-value) calculated using permutation analysis (see **Methods**). Thresholds were generated for representative traits (ALT and Red Blood Cell Count) across single-ancestry (Admixed American, African) and a pooled multi-ancestry cohort setting. The global study-wide median -log10 P-value threshold was determined to be 5.06, corresponding to $P = 8.7 \times 10^{-6}$.

#### Table S4: Admixture mapping associations using GNOMIX LAI labels

Detailed summary of independent region-trait associations meeting genome-wide significance. For each locus, the table provides the trait, trait category, phenotype, and genomic coordinates (hg19) and tested cohort. Association statistics (signed -log10 P-value and effect size) are reported for both NAT and AFR local ancestries to allow for direct comparison of effect sizes. "Local Ancestry Associated" indicates the specific ancestry driving the signal (passing the empirical significance threshold). "Trait Region Overlap" delineates pleiotropic regions associated with multiple phenotypes.

#### Table S5: Annotation of ADM associations using the GWAS Catalog

Mapping of significant local ancestry associations to known gene-phenotype associations. Genomic regions identified in the admixture mapping analysis were annotated with genes overlapping the top GNOMIX bin using GENCODE v44. These genes were then queried against the GWAS Catalog to identify previously reported trait associations. "GWAS Catalog Traits" lists all phenotypes currently associated with the mapped gene, allowing for the identification of known and novel gene-trait pairs where the specific phenotype in this study has not been previously reported for the associated gene.

#### Table S6: African local ancestry frequencies across single ancestry associations

Characterization of AFR local ancestry composition across phenotype-associated genomic intervals. Columns specify the trait and hg19 coordinates, while distinguishing the discovery cohort from the analysis cohort. The final column, "Freq_AFR", quantifies the aggregate frequency of AFR local ancestry for the indicated cohort at each locus.

#### Table S7: Genomic inflation factors across cohorts.

Assessment of test statistic inflation ($\lambda_{GC}$) and consistency of association signals. Columns display the genomic inflation factor ($\lambda_{GC}$) for admixture mapping in single-ancestry (Admixed American, African and pooled cohorts. "Abs Diff" quantifies the absolute difference in inflation in pooled vs. single-ancestry cohorts.
